# DeepFake electrocardiograms: the key for open science for artificial intelligence in medicine

**DOI:** 10.1101/2021.04.27.21256189

**Authors:** Vajira Thambawita, Jonas L. Isaksen, Steven A. Hicks, Jonas Ghouse, Gustav Ahlberg, Allan Linneberg, Niels Grarup, Christina Ellervik, Morten Salling Olesen, Torben Hansen, Claus Graff, Niels-Henrik Holstein-Rathlou, Inga Strümke, Hugo L. Hammer, Molly Maleckar, Pål Halvorsen, Michael A. Riegler, Jørgen K. Kanters

## Abstract

Recent global developments underscore the prominent role big data have in modern medical science. Privacy issues are a prevalent problem for collecting and sharing data between researchers. Synthetic data generated to represent real data carrying similar information and distribution may alleviate the privacy issue.

In this study, we present generative adversarial networks (GANs) capable of generating realistic synthetic DeepFake 12-lead 10-sec electrocardiograms (ECGs). We have developed and compare two methods, WaveGAN* and Pulse2Pulse GAN. We trained the GANs with 7,233 real normal ECG to produce 121,977 DeepFake normal ECGs. By verifying the ECGs using a commercial ECG interpretation program (MUSE 12SL, GE Healthcare), we demonstrate that the Pulse2Pulse GAN was superior to the WaveGAN to produce realistic ECGs. ECG intervals and amplitudes were similar between the DeepFake and real ECGs. These synthetic ECGs are fully anonymous and cannot be referred to any individual, hence they may be used freely. The synthetic dataset will be available as open access for researchers at OSF.io and the DeepFake generator available at the Python Package Index (PyPI) for generating synthetic ECGs.

In conclusion, we were able to generate realistic synthetic ECGs using adversarial neural networks on normal ECGs from two population studies, i.e., there by addressing the relevant privacy issues in medical datasets.

## Introduction

The use of artificial intelligence (AI) has increased in medicine over the past years. AI in medicine is to aid clinicians with decisions that are more accurate and to improve personalized medicine. The prerequisite and foundation for artificial intelligence is the large amount of high-quality clinical data.

With updates of the General Data Protection Regulation (GDPR) regulative in the EU, the free flow of data has been restricted to ensure patient consent and anonymity^1^. Even anonymized or deidentified data cannot be shared between research groups in different countries, because combining few variables in an anonymized dataset, may allow for individual identification^2^. For example, knowing the zip code, birthday and sex is enough to identify 87% of US citizens^3^. However, large-scale, publicly available open-access medical datasets are required for personalized medicine to improve data-heavy machine learning solutions in medicine.

Generating of realistic synthetic data is an alternative solution to the privacy issue. Synthetic data should contain all the desired characteristics of a specific population, but without any sensitive content, making it impossible to identify individuals. Therefore, properly generated synthetic data are a solution to the privacy problem and can enable data sharing between research groups.

An electrocardiogram (ECG) is a voltage time series reflecting the electric currents within the heart, a widely used, easy applicable and inexpensive clinical screening procedure to detect cardiac diseases. Using multiple electrodes, 3D propagation of cardiac electric impulses can be obtained and plotted as a standard 10-sec 12-lead ECG.

In this paper, we showcase synthetic ECGs as an example of complex medical data. Synthetic ECGs have been a topic of interest and research for many years. McSharry et al.^4^ and Sayadi et al.^5^ proposed mathematical dynamical models to generate continuous ECG signals, but these models were restricted to only one lead and did not reflect the distribution found in the normal population, nor did they give any insight in the mechanisms behind the disease.

Generative adversarial networks (GAN) were introduced in 2014 by Goodfellow et al. to generate synthetic data^6^ using multi-layer perceptrons. A GAN consists of two deep neural networks: A generator network making signals (here ECGs) from random noise and a discriminator network evaluating whether the ECG is real or fake. During training, a mix of real ECGs and generated DeepFake ECGs are presented to the discriminator, which assigns a score to the ECG. A high score represents a likely real ECG, and a low score a supposed DeepFake ECG. As training proceeds, both the generator and the discriminator improve until an equilibrium is reached^7^. Later, Radford et al.^8^ developed a convolutional GAN to generate synthetic images, which is well suited for images like the ECG.

Since ECGs basically are time series, our initial approach was to use a WaveGAN^9^ which is capable of generating sound signals. The classical WaveGAN is only able to output a single channel time series, so we modified the WaveGAN to generate all ECG channels (denoted WaveGAN*) instead of audio signals. We also introduced a novel DeepFake ECG U-net generative model, called Pulse2Pulse inspired by WaveGAN published by Donahue et al.^9^ and compare our Pulse2Pulse generator to the WaveGAN generator.

In this paper, we present two GANs with the ability to generate an unlimited number of 10-sec 12-leads synthetic “DeepFake” ECGs as a solution to overcome privacy issues related to real ECG data. These DeepFake ECGs can be openly distributed and freely downloaded as open access to be used by other scientists to develop ECG algorithms.

## Results

We used ECGs from two population studies (GESUS^10^ and Inter99^11^). To avoid chimeras between normal and abnormal ECG, we only trained the neural network with ECGs classified as normal by the MUSE 12SL. As shown in Table 1, both the WaveGAN* and Pulse2Pulse improved during training expressed as the percentage of DeepFake ECGs classified by the MUSE 12SLas normal ECGs. The Pulse2Pulse GAN trained faster than the WaveGAN* and had a better performance (expressed as fraction of ECGs classified as normal by the MUSE) than the WaveGAN* at their respective optimal number of training epochs (Table 1). Figure 1 shows a comparison of real and DeepFake ECGs, and the supplementary Figure S1 shows twenty randomly chosen DeepFake ECGs. Figure 2 shows the distribution of heart rates in the DeepFakes. By clinical definition Normal ECGs heart rates are between 60 and 99 beats per minute. The MUSE 12SL^12^ classified 129 DeepFakes (0.5%) as sinus tachycardia (fast heart rate≥100) and 2863 (10.2%) as sinus bradycardia (slow heart rate<60). Figure 4 shows that cross correlation between as an example the QT interval and the RR interval were preserved. All covariance structures can be seen in Supplementary Figure S2.

**Table 1.** Quantitative difference between WaveGAN* and Pulse2Pulse GAN in the initial training for determining the optimal network and optimal number of epochs. The best values are bolded for each GAN.

| Checkpoint (epochs) | Fraction of DeepFake ECGs classified as Normal (%) |  |
| --- | --- | --- |
|  | WaveGAN* | Pulse2Pulse |
| 500 | 20.9 | 78.7 |
| 1000 | 69.5 | 81.2 |
| 1500 | 71.2 | 78.8 |
| 2000 | **72.5** | 79.7 |
| 2500 | 71.3 | **81.6** |
| 3000 | 65.3 | 81.5 |

**Figure 1.**
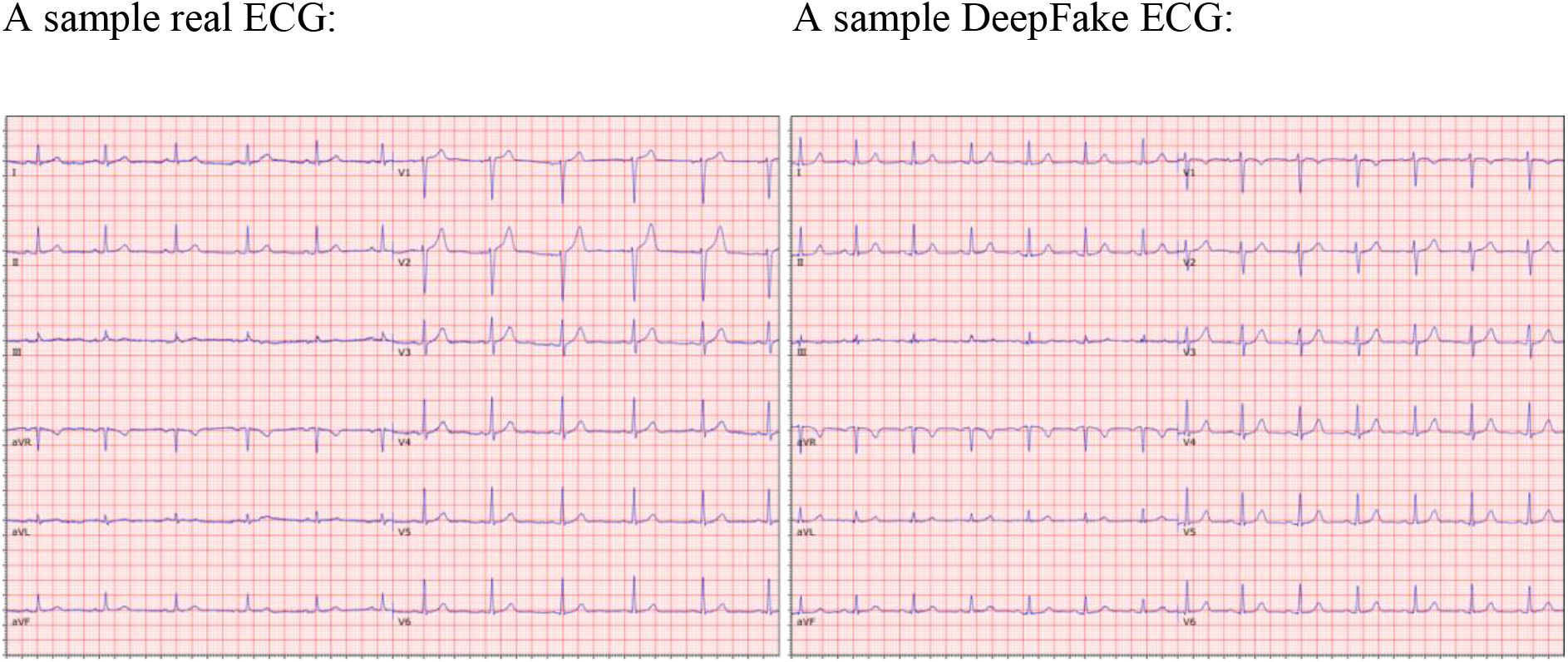
Comparison of examples of a real ECG (left lane) and a DeepFake ECG (right lane). See supplementary Figure S1 for 20 more randomly chosen pairs of real and DeepFake ECGs.

**Figure 2.**
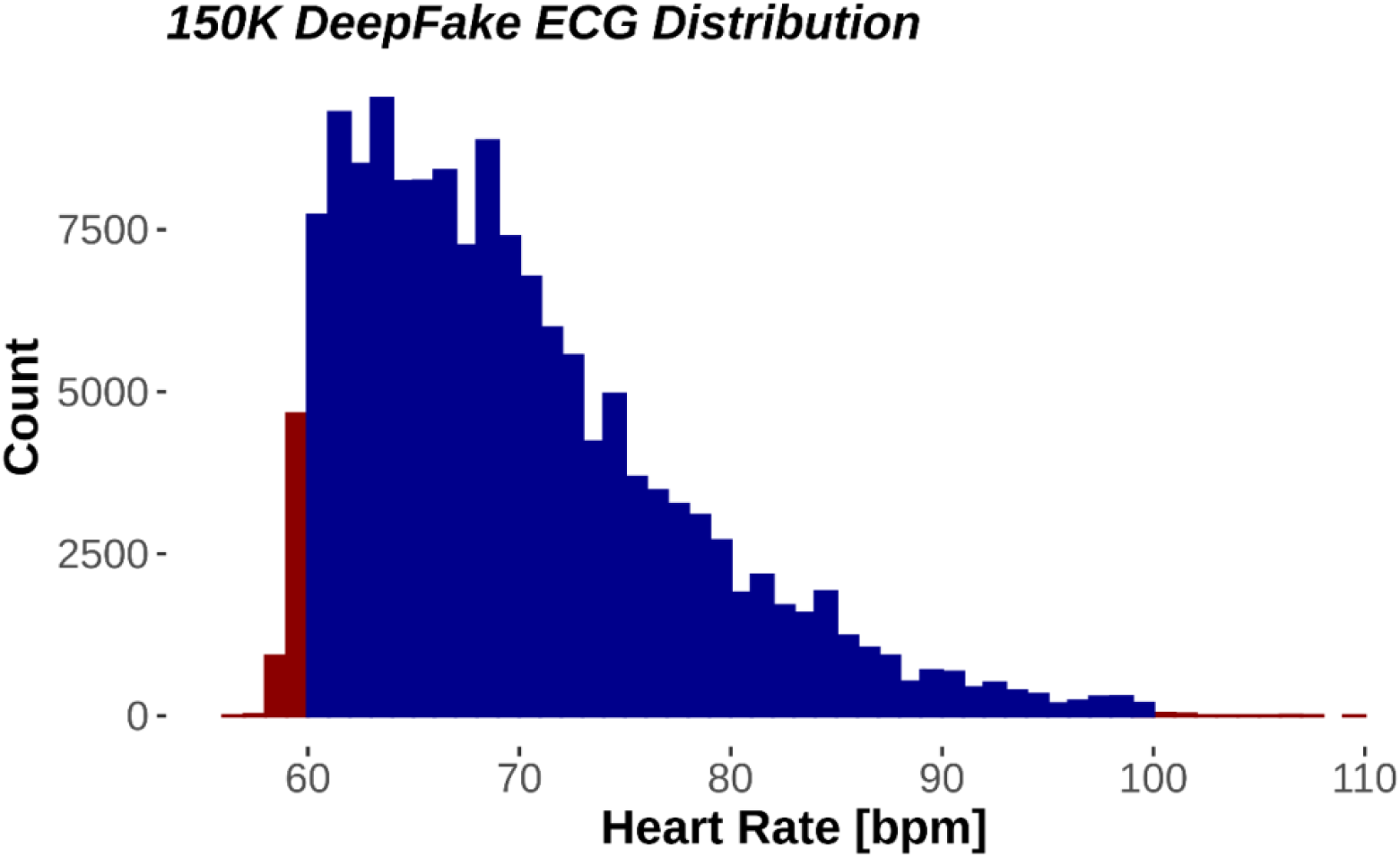
Distribution of heart rates in all 150.000 DeepFake electrocardiograms. Red fill denotes outside the normal heart rate range. Blue fill is within normal heart rate range (60-100).

The generated DeepFake ECGs can be downloaded at OSF.io (https://osf.io/6hved/) with the corresponding ground truth parameters for the QT, RR, PR and QRS intervals and the P, STJ, R, and T amplitudes (see Figure 3 for ECG wave/interval naming terminology) delivered by the MUSE 12SL system (version 2.43). The DeepFake ECGs may be freely used for scientific use or commercial algorithm development if this paper is properly cited.)

**Figure 3.**
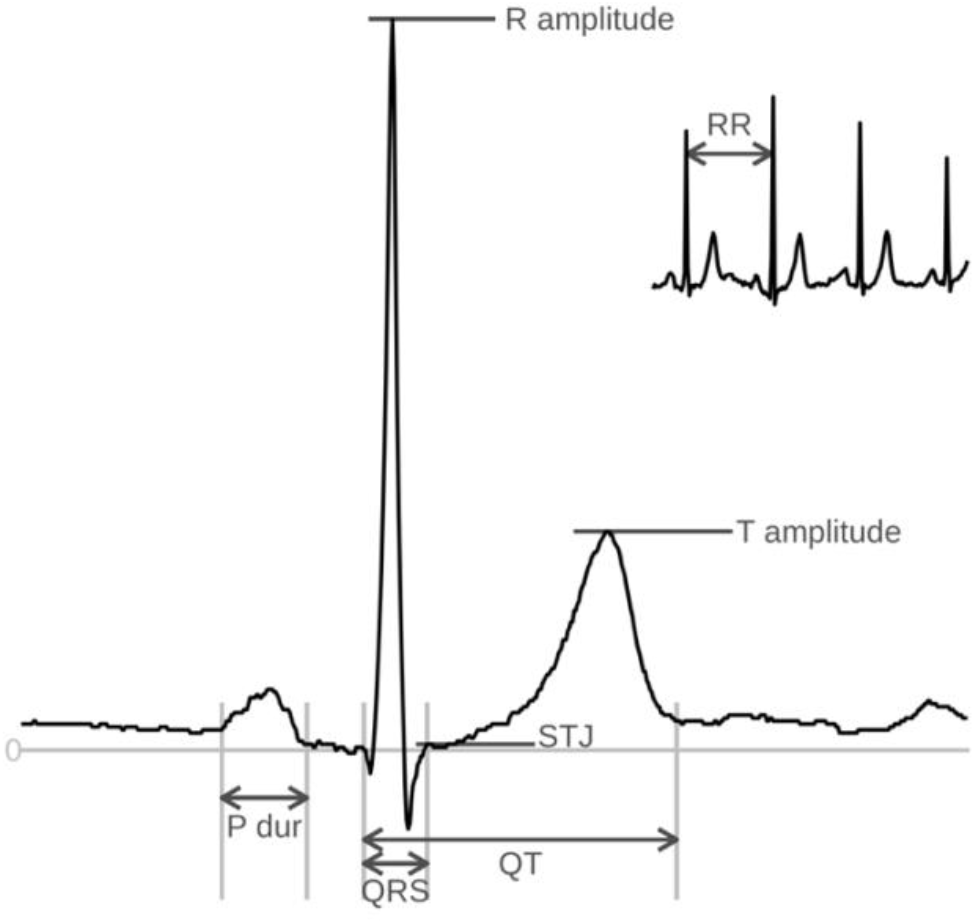
An ECG complex with the nomenclature of intervals (QT, QRS, P duration) and Amplitudes (STJ, R, T) and RR-interval (which can be converted to heart rate (HR) as HR=60/RR interval.

**Figure 4.**
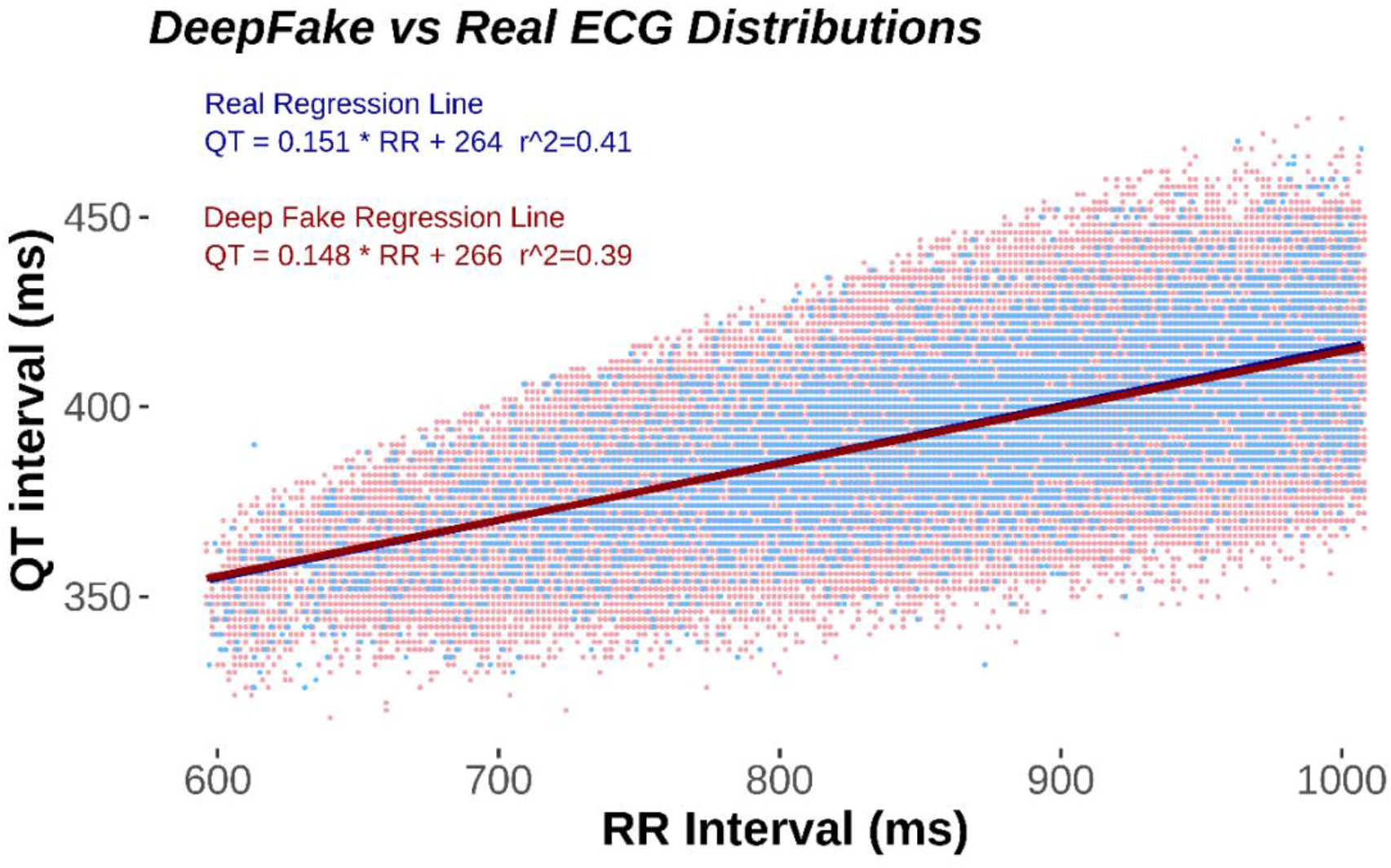
Scatter plot of the QT/RR interval relationship. Real ECG in blue and normal DeepFakes in red. DeepFake dots are nudged 1 ms to the left for visibility. Note that there are 121,977 normal DeepFakes and only 7,233 Real ECG making the DeepFake distribution more pronounced. As seen by the correlation coefficient r^2^, the real and the fake DeepFake ECGs are similarly distributed.

Using the Pulse2Pulse model from the optimal number of epochs (2500), we generated 150,000 DeepFake ECGs. To ensure that these ECGs were realistic, we uploaded the 150,000 ECGs to the GE MUSE system and analyzed them using the 12SL algorithm. We found that 81.3% of the 150,000 DeepFake ECG were classified as “Normal ECG” (vs. 81.6 % in the initial training). Table 2 compares real vs. DeepFake ECGs using eight ECG properties (heart rate, P duration, QT interval, QRS duration, PR interval, STJ amplitude, R amplitude, and T amplitude extracted using MUSE 12SL. See Figure 3 for ECG nomenclature). The real data included all ECGs from GESUS and Inter99 classified as “Normal ECG” which were used for training. DeepFake ECGs are presented both as all 150.000 generated ECGs and the subset classified as Normal ECG. The supplementary Table S4 summaries the most common reasons for classifying DeepFake ECGs as Non-Normal ECGs.

**Table 2.** Mean, standard deviation (std), 2.5%, and 97.5% percentile for standard ECG parameters in real and fake ECGs. BPM = beats per minute

|  |  | Real – Normal (7,233) |  |  |  | Pulse2Pulse – Normal (121,977) |  |  |  | Pulse2Pulse – All (150,000) |  |  |  |
| --- | --- | --- | --- | --- | --- | --- | --- | --- | --- | --- | --- | --- | --- |
|  |  | Mean | std | 2.5% | 97.5% | Mean | std | 2.5% | 97.5% | Mean | std | 2.5% | 97.5% |
| Heart rate | BPM | 70 | 8 | 60 | 90 | 70 | 7 | 60 | 88 | 70 | 8 | 60 | 89 |
| P Duration | ms | 105 | 12 | 82 | 130 | 117 | 17 | 86 | 152 | 118 | 17 | 84 | 152 |
| QT Interval | ms | 395 | 21 | 352 | 436 | 395 | 20 | 354 | 436 | 395 | 22 | 352 | 436 |
| QRS Duration | ms | 90 | 9 | 74 | 110 | 92 | 9 | 78 | 112 | 93 | 10 | 78 | 114 |
| PR Interval | ms | 156 | 19 | 120 | 198 | 158 | 17 | 126 | 192 | 159 | 19 | 124 | 194 |
| STJ amplitude (V5) | μV | 2 | 27 | -44 | 58 | 18 | 33 | -44 | 87 | 16 | 36 | -54 | 87 |
| R Amplitude (V5) | μV | 1287 | 402 | 600 | 2163 | 1275 | 367 | 620 | 2026 | 1273 | 402 | 566 | 2094 |
| T Amplitude (V5) | μV | 343 | 137 | 126 | 664 | 366 | 135 | 156 | 668 | 361 | 141 | 141 | 673 |

## Discussion

Although deep learning has been previously used for ECG analysis^13,14^, this study is the first study to generate realistic synthetic 10-sec 12-lead DeepFake ECGs. We demonstrate that the characteristics from the real ECGs were similar to DeepFake ECGs. Hence, our DeepFake generator was able to construct synthesized ECG with similar intervals and amplitudes as the original population.

In our study, nearly one fifth of the DeepFake ECGs were not recognized as Normal ECGs (Non-Normal) by the commercial MUSE 12SL ECG analyzer (No ECGs were rejected as being invalid). Many ECG parameters use hard boundaries in distinguishing between Normal and Non-Normal. For example, a normal heart rate is defined as between 60 to 99 bpm. Since we trained our model only on Normal ECGs, the input distribution for the GAN was a truncated asymmetric distribution. Thus, the clinically defined boundaries are skewed compared to the normal distribution of heart rates. The left truncation (at low heart rates) will discard more individuals than the right truncation (at high heart rates), and the final distribution of the real ECGs will be close to a truncated normal distribution with asymmetric truncations. The GAN will generally learn that heart rates outside 60-99 are not valid, but small deviations will occur as seen in Figure 2 and Table 2. Since similar boundaries exist for many ECG parameters (for example PR interval >120 ms or QRS Interval<120 ms) sharp truncations occur with several ECG parameters. This could lead to exclusion of some DeepFake ECGs, simply because the ECG intervals or amplitudes were just outside the normal range. Most ECG amplitudes and intervals were similar between real ECGs and DeepFake ECGs. It is noteworthy that the STJ amplitude and the P duration had the greatest deviation between real ECGs and DeepFake ECGs. This may be due to both STJ and P amplitudes are small, and that the network may tend to focus on larger waves such as the R and T waves. Following this theory, the network would to some extent neglect the smaller waves and features thereby introducing a larger uncertainty. Future networks may improve the ECG generation using conditional GANs to give more attention to smaller signal features. The Pulse2Pulse model was able to preserve the covariance structure between different ECG features, as seen in the most important relationship the QT/RR relationship which is known to have prognostic importance^15^.

A challenging task is to define the optimal number of epochs for training. GANs tend to become unstable during the training process with the risk of the generator producing unrealistic output. To get an unbiased estimate on how well the trained GAN performs, we used the commercial MUSE 12SL system which automatically and reliably evaluates an ECG with a sensitivity of 99.9% and specificity of 100%^16^. Although the ECG discarded by the MUSE 12 SL may only have minimal abnormalities (like a heart rate of 59.9 where 60 is normal), the filtering of the DeepFake ECGs ensures, that the best epoch is chosen without bias, and the resulting ECGs are normal not only according to the discriminator, but also according to one of the most widely used ECG system in hospitals worldwide.

Personalized medicine is dependent on big data, which frequently is facilitated by international cooperation to ensure large datasets for both researchers and industry. However, privacy and general data protection regulation rules are major obstacles for sharing data between researchers from different institutions and countries, or with the industry^17^.

In conclusion, by constructing synthetic signals from real patients keeping the same clinical information as in the real dataset, we show how to overcome privacy and ethical^18^ concerns for data sharing. The synthetic data generated by our Pulse2Pulse GAN makes it impossible to identify any patients, but still the ECGs remain useful for data scientists and the industry to use for developing novel algorithms for ECG analysis. The approach is not limited to ECGs but could be generalized to all medical multichannel data, e.g., electroencephalography and electromyography. Therefore, the DeepFake ECGs generated from the Pulse2Pulse model can be used as a replacement to overcome the privacy constraints in real datasets.

## Methods

The WaveGAN model is an evolution from the first GAN model introduced by Goodfellow et al.6. The two deep neural networks named generator (G) and discriminator (D) to achieve the generation task. The main goal of the generator is to produce a data sample input (ECG(z)) from random noise (z) to present for the discriminator. The discriminator’s task is to differentiate between real and fake data. We train the generator and discriminator together as a competition (minmax game) between them. When a steady state is reached, the training halts, and the generator will generate realistic synthetic ECGs.

### Data preparation

We used two combined datasets: the Danish General Suburban Population Study^10^ (GESUS) and the Inter99 study^11^ (CT00289237, ClinicalTrials.gov). GESUS consists of 8,939 free-living subjects, and Inter99 consists of 6,667 free-living subjects with an available digital ECG. To avoid generation of hybrid ECG with mixed ECG abnormalities not occurring in real persons (e.g., to both be in sinus rhythm and atrial fibrillation at the same time which is impossible), we excluded ECGs who were not classified as normal (n=8,348) leaving 7,233 Normal ECGs for training.

A 12-lead 10-sec ECG consists only of 8 independent channels since 4 of the channels are simply trigonometric rotations. Therefore, the input ECG signal is 5,000×8 data points (corresponding to 10 sec with 500 samples per sec. x 8 channels). In addition to the up-scaling, we calculate the missing four channels with trigonometric functions to create the classic 12-channels ECG.

#### WaveGAN*

The input to WaveGAN* is a 1D random noise vector sampled from the uniform distribution (mean = 0, std = 1) with 100 x 1 passes through six deconvolution blocks to generate the desired output of 5000 x 8 samples. The deconvolution blocks are built from a series of four layers: an up-sampling layer, a constant padding layer, a 1D-convolution layer, and a ReLU activation function, consecutively. This implementation is deeper than the original architecture which use five deconvolution blocks used to generate synthetic music samples. Table S1 has comprehensive details of our WaveGAN* generator network.

#### Pulse2Pulse

The implementation of the Pulse2Pulse architecture (Figure 4) is inspired by the U-Net architecture^19^ which is used for image segmentation. However, our Pulse2Pulse implementation is different than the original U-net implementation because the Pulse2Pulse implementation use 1D CNN for ECG signal generation rather than the 2D CNN used for original image segmentation task. The Pulse2Pulse network takes an 8×5000 noise vector which has the same dimension as the output dimension of a generated ECG. Then, we pass the noise through six down-sampling blocks followed by six up-sampling blocks as illustrated in Figure 3b. Each down-sampling block consists of a 1D-convolution layer followed by a Leaky ReLU activation. The up-sampling block is similar the deconvolution block used in WaveGAN*. In down-sampling, we have used Leaky ReLU instead of the ReLU layer used in the up-sampling to match the down-sampling operations to the discriminator. In addition to the up-sampling and down-sampling, the major modification is a bypass with down-sampling block features concatenating into the up-sampling block features represented by the black arrows in Figure 3b. To facilitate for this concatenation, we doubled the input size of up sampling blocks compared to WaveGAN* up sampling blocks. More details about Pulse2Pulse architecture are shown in the supplementary Table S1.

#### Discriminator

The same discriminator is used by WaveGAN* and Pulse2Pulse to discriminate between real and fake ECGs (Figure 3c). We used seven convolution layers (the original WaveGAN^9^ has five layers), and each convolution layer is followed by a Leaky ReLU activation and the phase shuffle layer introduced in the original WaveGAN paper^9^. The discriminator takes an ECG as input (5000 samples * 8 channels) and outputs a score how close the ECG are to be determined fake or real.

**Figure 4.**
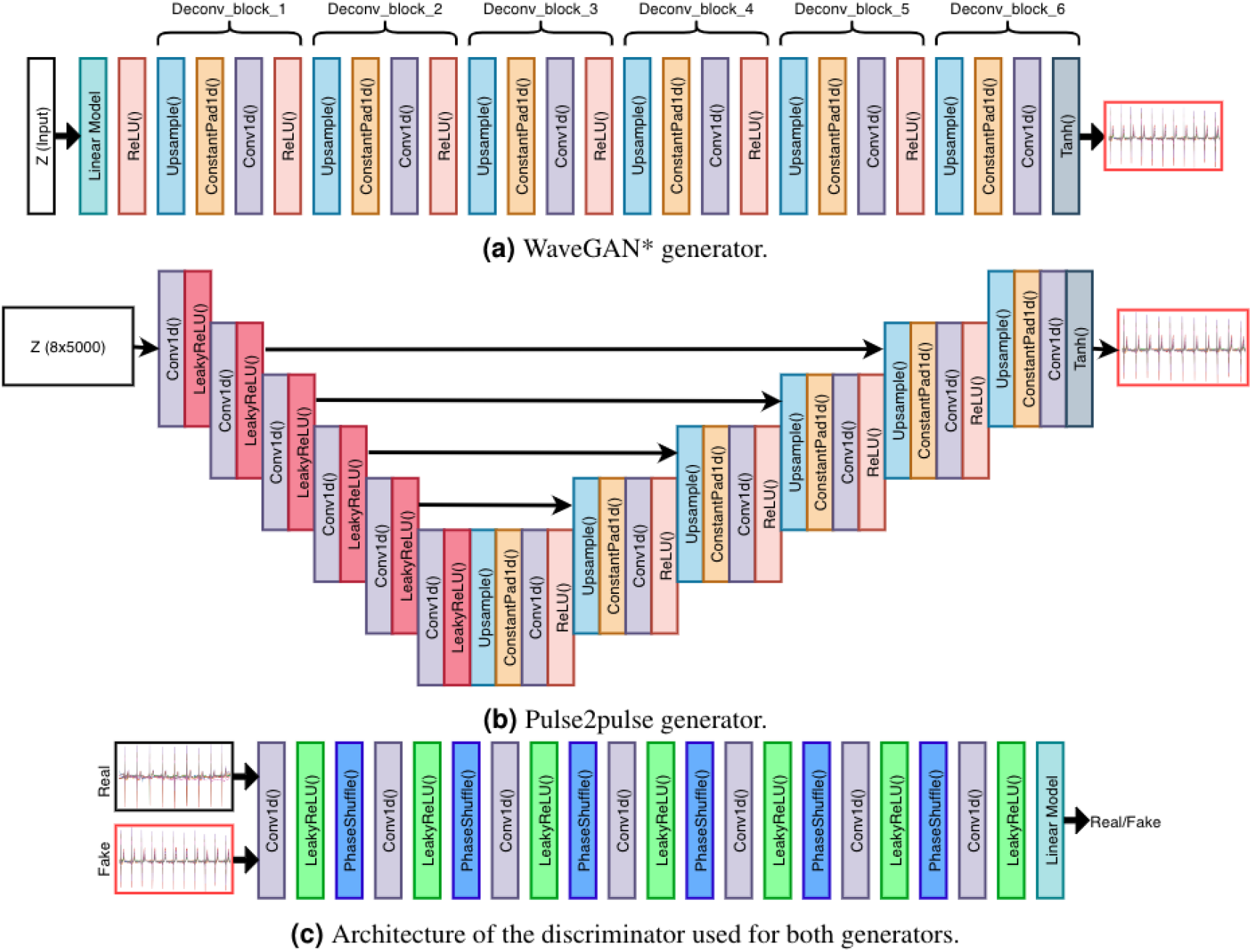
Model architectures of the generators and the discriminator used to generate synthetic ECGs. WaveGAN* uses a 1D noise vector with 100 points. Pulse2Pulse uses a 2D noise vector with size of 8×5000 as input, same as the output ECG size.

#### Training

The models were trained on a Ubuntu workstation with a double Xeon processor and a GeForce NVIDIA RTX 2080 running the Pytorch deep learning framework^20^. We ran all our experiments (generators + discriminator) using the Adam^21^ optimizer with a learning rate of 0.0001, β1value of 0.5, and β2 value of 0.9. As loss function, we used gradient clipping WGAN-GP^22^, to ensure faster and better convergence. Similar to the audio generation paper of WaveGAN^9^, we updated (backpropagated) the discriminator five times per update of the generator. We used a batch size of 32, which is half of the original batch size of 64 used in the original WaveGAN paper, as a result of using larger networks than the WaveGAN networks. We kept the training process until 3000 epochs (∼10 days computing time) because we experienced unstable training curves for both WaveGAN* and Pulse2Pulse afterwards.

#### DeepFake ECGs

For evaluation of our two GAN models, we initially generated 10,000 ECGs from every 500 epochs until 3000 epochs from each GAN model. The DeepFake ECGs were transferred to the MUSE system and evaluated by the MUSE 12SL algorithm v. 2.43^12^ using the fraction of DeepFake ECGs described as Normal (similar to the Real ECGs used for training). Using the best epoch for the best GAN, we generated 150.000 DeepFake ECGs. These DeepFakes were similar evaluated by the MUSE 12SL.

## Data Availability

The Normal DeepFake ECGs are available at OSF (https://osf.io/6hved/) with corresponding MUSE 12SL ground truth values freely downloadable and usable for ECG algorithm development. The DeepFake generative model is available at https://pypi.org/project/deepfake-ecg/ to generate only synthetic ECGs.

https://pypi.org/project/deepfake-ecg/

## Code Availability

The complete source code of all networks discussed in paper are available at GitHub (https://github.com/vlbthambawita/deepfake-ecg).

