## Supplementary material for "DeepFake electrocardiograms: the key for open science for artificial intelligence in medicine"

**Supplementary materials**

**Table S1**. Full details of the WaveGAN* Generator network. z = input noise vector, model dimension (d) = 50, batch size (n) = 32, channels (c) = 8.

| **Operation** | **Kernel** | **Size** |
|---|---|---|
| Input z | | (n,100) |
| Dense 1 | (100,10d) | (n,10d) |
| Reshape | | (n,5d,2) |
| ReLU | | (n,5d,2) |
| Deconv_block_1 (stride=1,upsample=5) | (25,5d,5d) | (n,5d,10) |
| Deconv_block_2 (stride=1,upsample=5) | (25,5d,3d) | (n,3d,50) |
| Deconv_block_3 (stride=1,upsample=5) | (25,3d,d) | (n,d,250) |
| Deconv_block_4 (stride=1,upsample=2) | (25,d,d/2) | (n,d/2,500) |
| Deconv_block_5 (stride=1,upsample=5) | (25,d/2,d/5) | (n,d/5,2500) |
| Deconv_block_6 (stride=1,upsample=2) | (25,d/5,c) | (n,c,5000) |

**Table S2**. Full details of the Generator network of Pulse2Pulse. z = noise vector (pseudo ECG), model dimension (d) = 50,batch size (n) = 32, channels (c) = 8. Deconvolution layers with * mark take output features from the 2nd, 3rd, 4th, and 5th convolutional layers and concatenate into channel dimension.

| Operation | Kernel | Size |
|---|---|---|
| Input z | | (n,c,5000) |
| Conv1D (Stride=2) | (25,c,d/5) | (n,d/5,2500) |
| LReLU (α=0.01) | | (n,d/5,2500) |
| Conv1D (Stride=5) | (25,d/5,d/2) | (n,d/2,500) |
| LReLU (α=0.01) | | (n,d/2,500) |
| Conv1D (Stride=2) | (25,d/2,d) | (n,d,250) |
| LReLU (α=0.01) | | (n,d,250) |
| Conv1D (Stride=5) | (25,d,3d) | (n,3d,50) |
| LReLU (α=0.01) | | (n,3d,50) |
| Conv1D (Stride=5) | (25,3d,5d) | (n,5d,10) |
| LReLU (α=0.01) | | (n,5d,10) |
| Conv1D (Stride=5) | (25,5d,5d) | (n,5d,2) |
| LReLU (α=0.01) | | (n,5d,2) |
| Deconv_block_1 (stride=1,upsample=5) | (25,5d,5d) | (n,5d,10) |
| Deconv_block_2* (stride=1,upsample=5) | (25,5dx2,3d) | (n,3d,50) |
| Deconv_block_3* (stride=1,upsample=5) | (25,3dx2,d) | (n,d,250) |
| Deconv_block_4* (stride=1,upsample=2) | (25,dx2,d/2) | (n,d/2,500) |
| Deconv_block_5* (stride=1,upsample=5) | (25,(d/2)x2,d/5) | (n,d/5,2500) |
| Deconv_block_6 (stride=1,upsample=2) | (25,d/5,c) | (n,c,5000) |

**Table S3**. Details of the discriminator network. Model dimension (d) = 50, channels (c) = 8, batch size (n) = 32.

| Operation | Kernel Size (width,channels,filters) | Output Shape |
|---|---|---|
| Input x or G(z) | | (n,c,5000) |
| Conv1D (Stride=2) | (25,c,d) | (n,d,2499) |
| LReLU (α=0.2) | | (n,d,2499) |
| Phase Shuffle (n=2) | | (n,d,2499) |
| Conv1D (Stride=2) | (25,d,2d) | (n,2d,1249) |
| LReLU (α=0.2) | | (n,2d,1249) |
| Phase Shuffle (n=2) | | (n,2d,1249) |
| Conv1D (Stride=2) | (25,2d,5d) | (n,5d,624) |
| LReLU (α=0.2) | | (n,5d,624) |
| Phase Shuffle (n=2) | | (n,5d,624) |
| Conv1D (Stride=2) | (25,5d,10d) | (n,10d,311) |
| LReLU (α=0.2) | | (n,10d,311) |
| Phase Shuffle (n=2) | | (n,10d,311) |
| Conv1D (Stride=2) | (25,10d,20d) | (n,20d,78) |
| LReLU (α=0.2) | | (n,20d,78) |
| Phase Shuffle (n=2) | | (n,20d,78) |
| Conv1D (Stride=2) | (25,20d,25d) | (n,25d,19) |
| LReLU (α=0.2) | | (n,25d,19) |
| Phase Shuffle (n=2) | | (n,25d,19) |
| Conv1D (Stride=2) | (25,25d,100d) | (n,100d,5) |
| LReLU (α=0.2) | | (n,100d,5) |
| Linear | | (n,25000,1) |

**Table S4**. MUSE 12SL diagnoses for the Non-Normal DeepFake ECGs. Note that an ECG may have several ECG diagnoses. The table shows only diagnoses with at least 250 occurrences.

| **Total number of Non-Normal DeepFake ECGs** | **28023** |
|---|---|
| Non-Specific ST abnormality | 7176 |
| Sinus Bradycardia | 2863 |
| Inferior Infarct | 2540 |
| Left Ventricular Hypertrophy | 1777 |
| Non-Specific intraventricular delay | 1610 |
| Low Voltage QRS | 1566 |
| Fusion complexes | 1244 |
| Anterior Infarct | 1238 |
| ACUTE MI/STEMI | 1003 |
| Left Axis Deviation | 826 |
| Lateral Infarct | 773 |
| Premature Ventricular complexes | 748 |
| Septal Infarct | 745 |
| Undetermined Rhythm | 630 |
| Premature Atrial Complexes | 615 |
| With short PR | 614 |
| With 1 degree AV block | 584 |
| Non-specific T wave abnormality | 526 |
| Prolonged QT | 460 |
| Non-specific interventricular block | 444 |
| ST elevation (inferior injury) | 405 |
| Rightward axis | 389 |
| Aberrant conduction | 384 |
| Left Ventricular Hypertrophy | 318 |
| Moderate Left Ventricular Hypertrophy | 310 |
| Incomplete Right Bundle Branch Block | 289 |
| Left atrial enlargement | 268 |

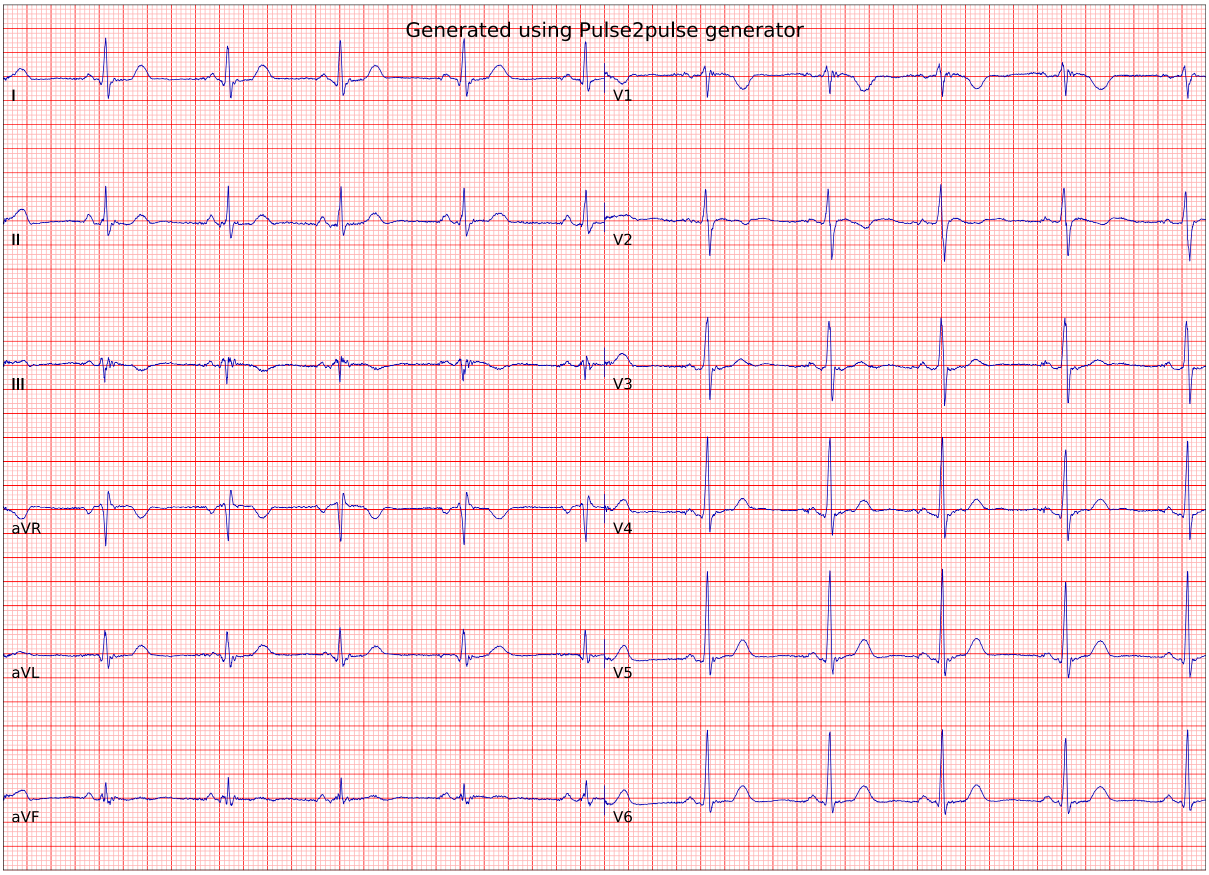
Generated using Pulse2pulse generator
I
II
III
aVR
aVL
aVF
V1
V2
V3
V4
V5
V6

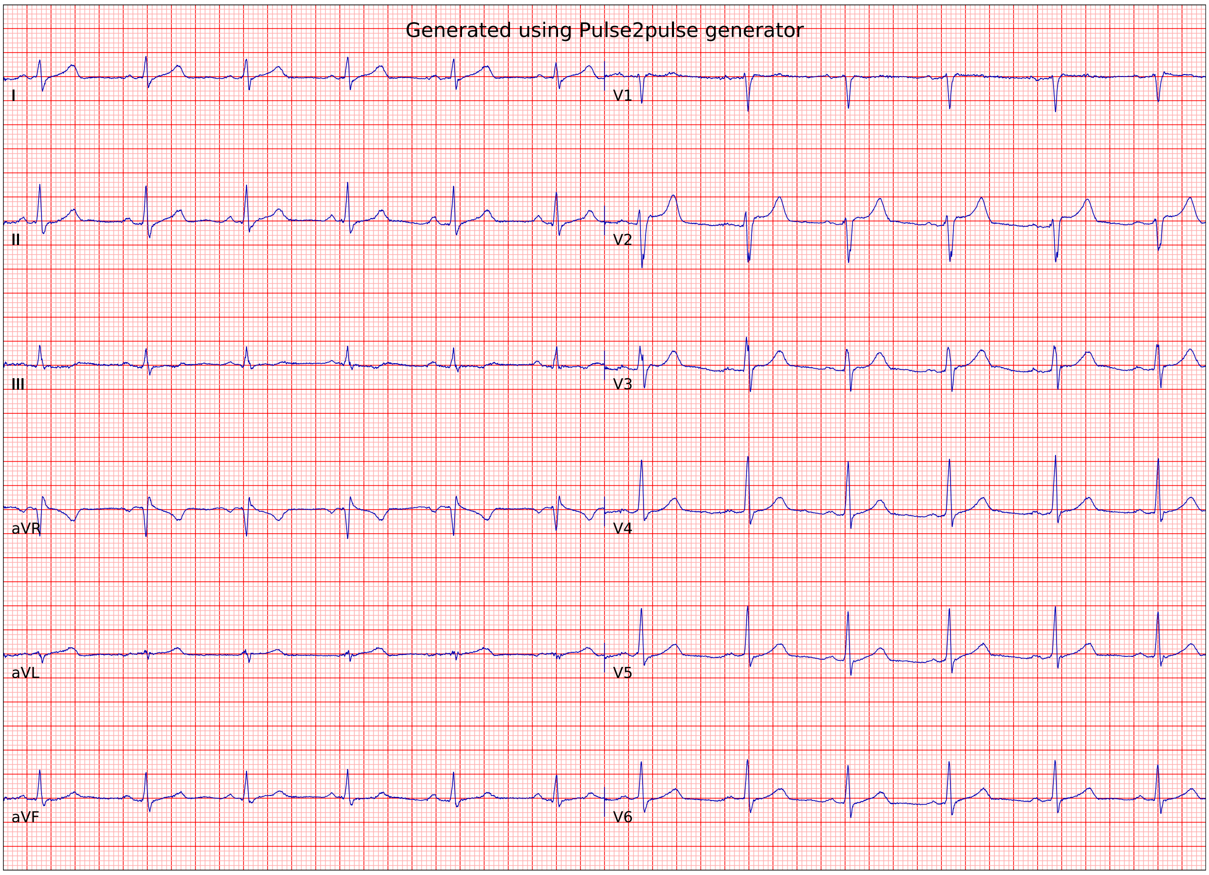
Generated using Pulse2pulse generator
I
II
III
aVR
aVL
aVF
V1
V2
V3
V4
V5
V6

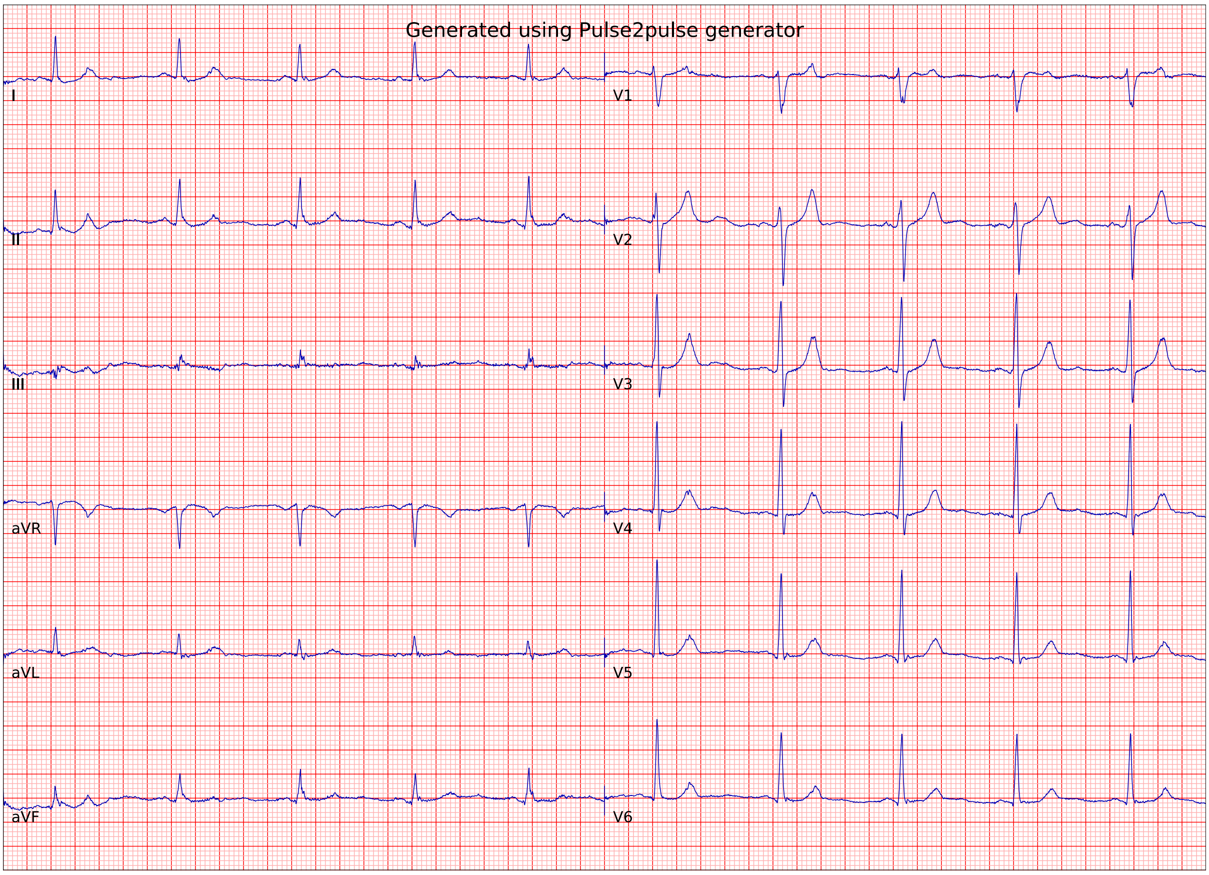
Generated using Pulse2pulse generator
I
II
III
aVR
aVL
aVF
V1
V2
V3
V4
V5
V6

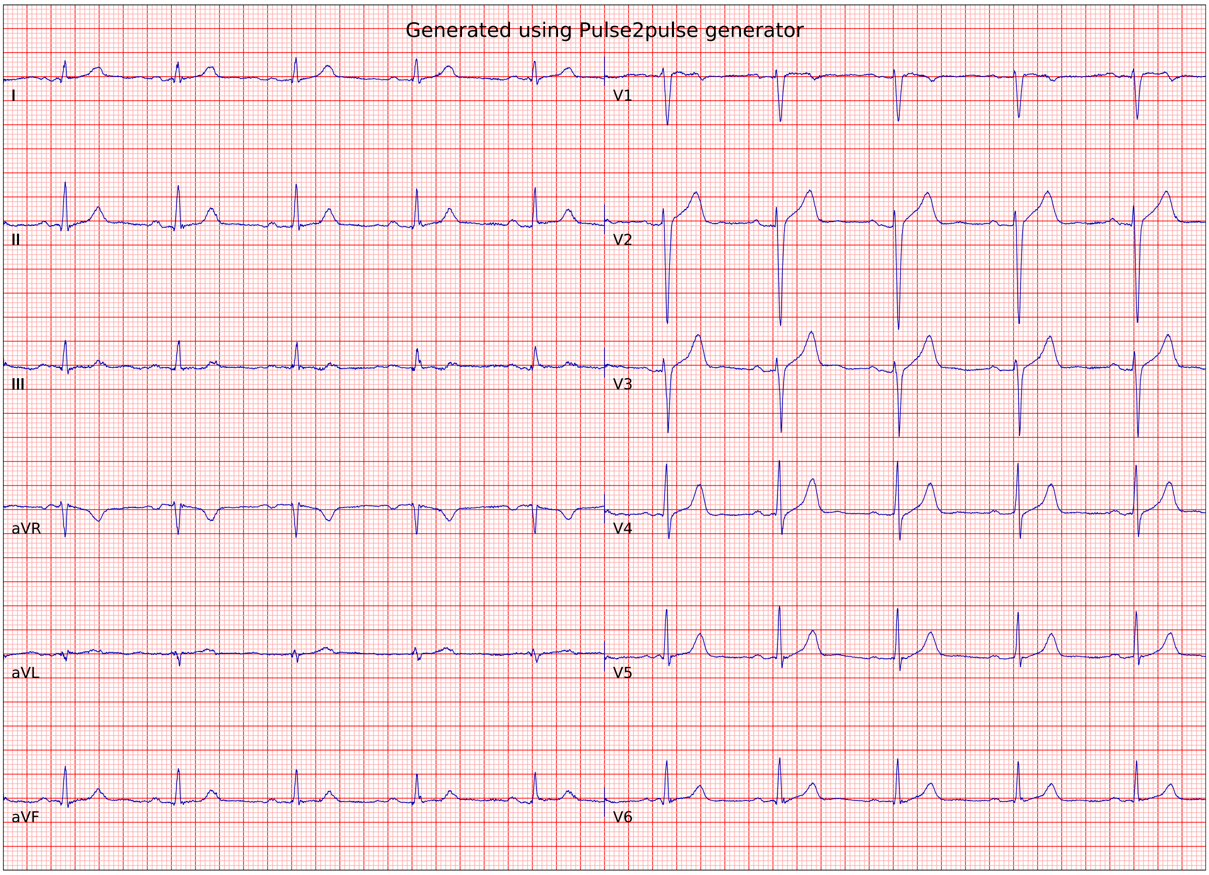
Generated using Pulse2pulse generator
I
II
III
aVR
aVL
aVF
V1
V2
V3
V4
V5
V6

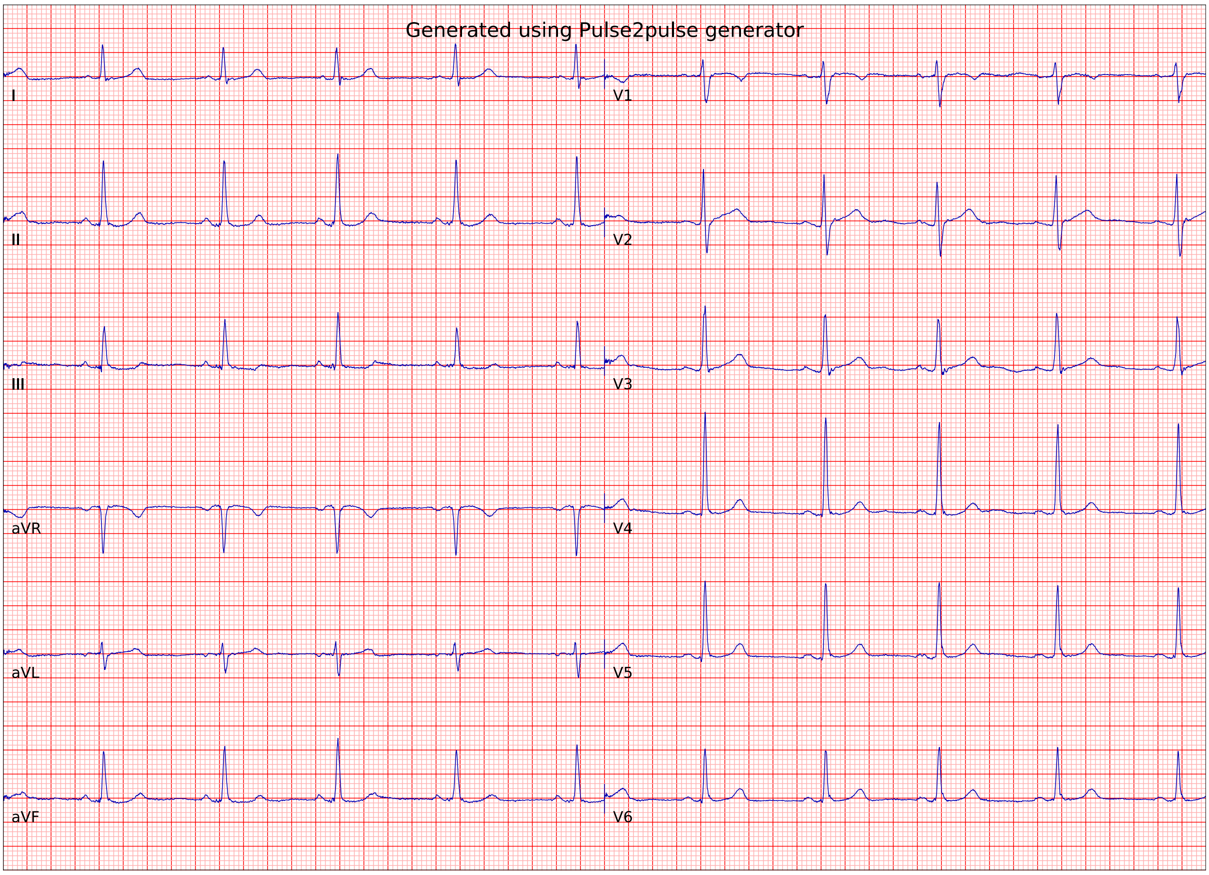
Generated using Pulse2pulse generator
I
V1
II
V2
III
V3
aVR
V4
aVL
V5
aVF
V6

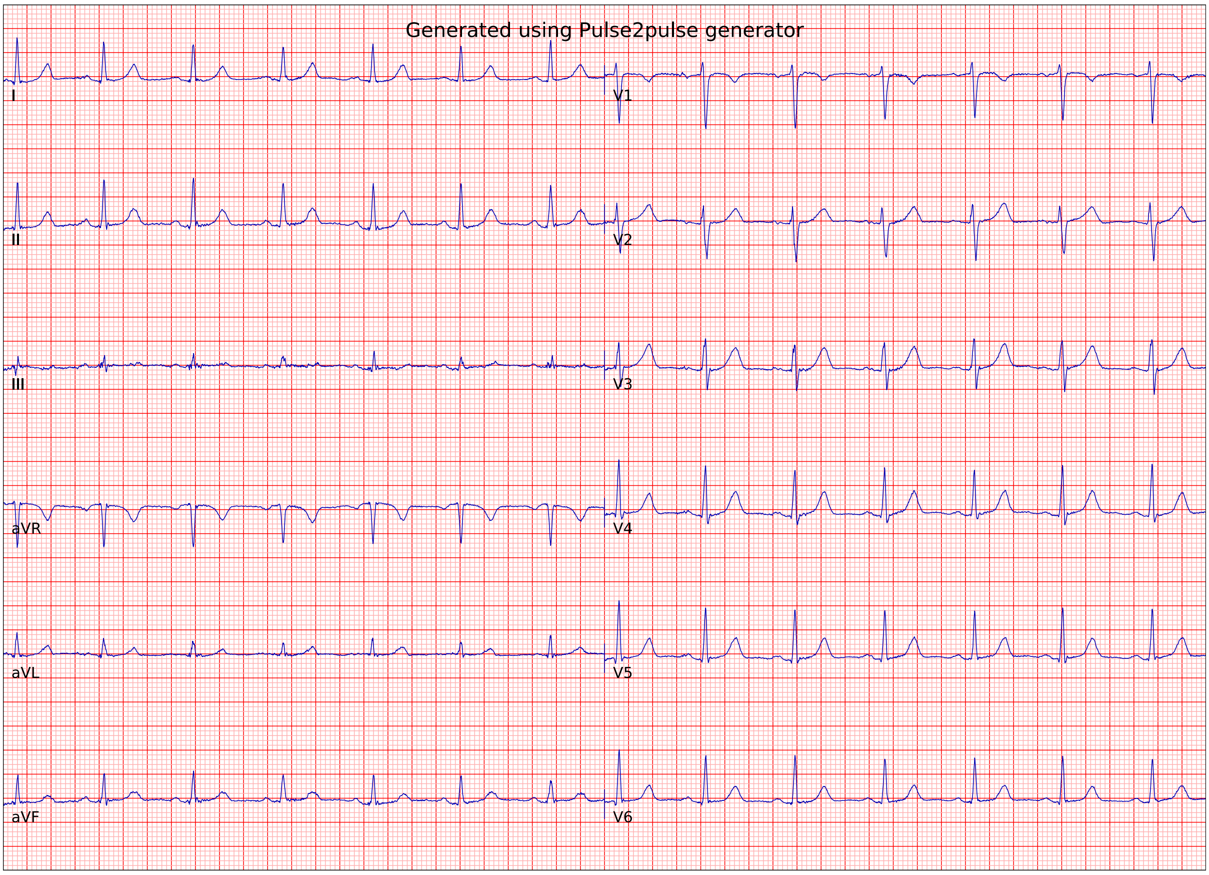
Generated using Pulse2pulse generator
I
V1
II
V2
III
V3
aVR
V4
aVL
V5
aVF
V6

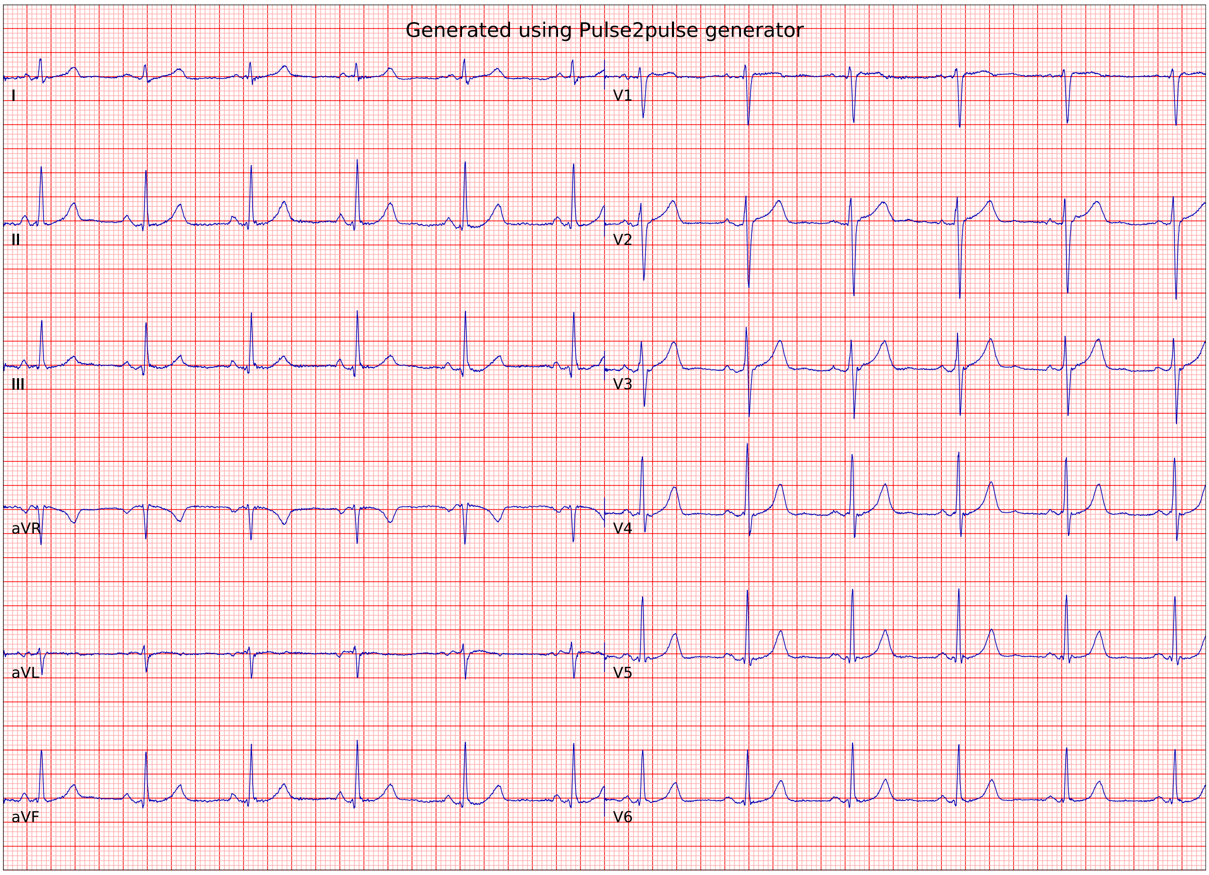
Generated using Pulse2pulse generator
I
II
III
aVR
aVL
aVF
V1
V2
V3
V4
V5
V6

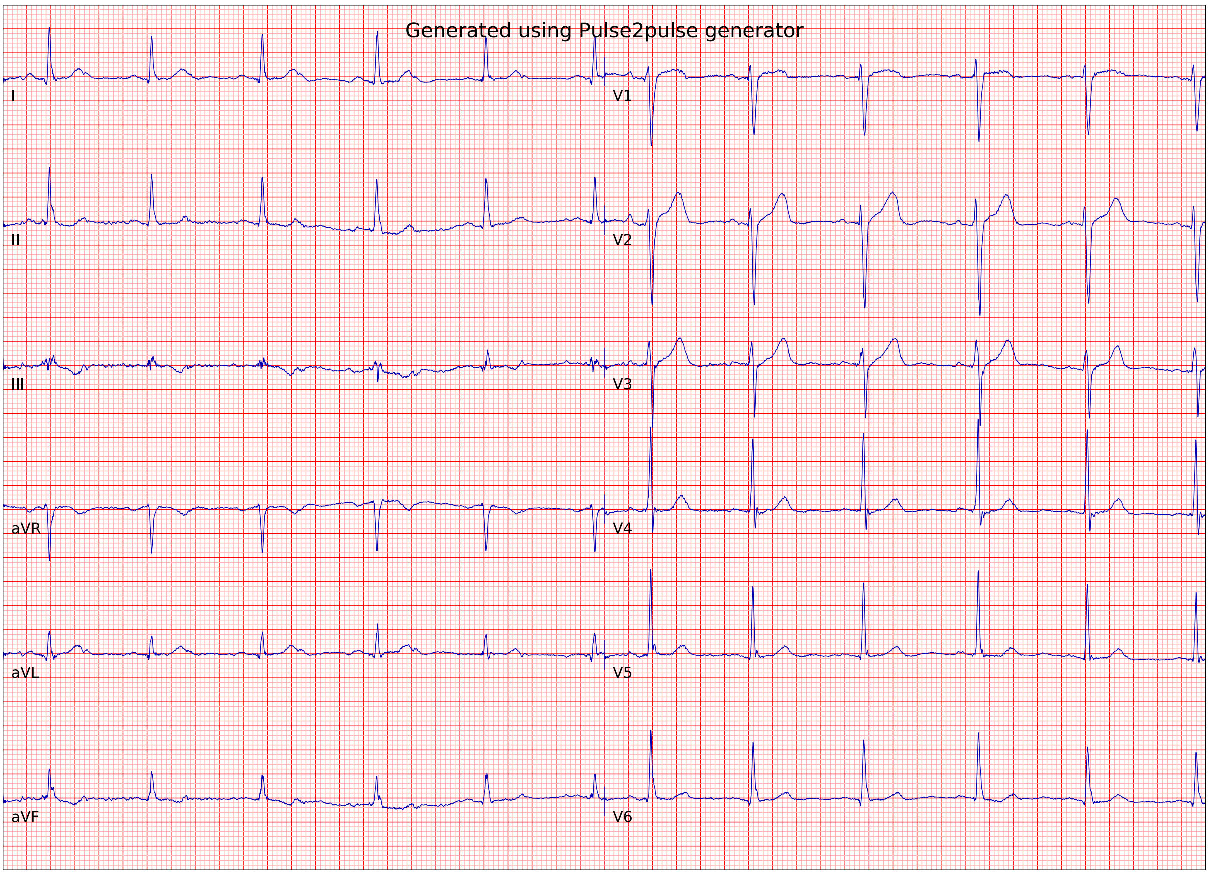
Generated using Pulse2pulse generator
I
II
III
aVR
aVL
aVF
V1
V2
V3
V4
V5
V6

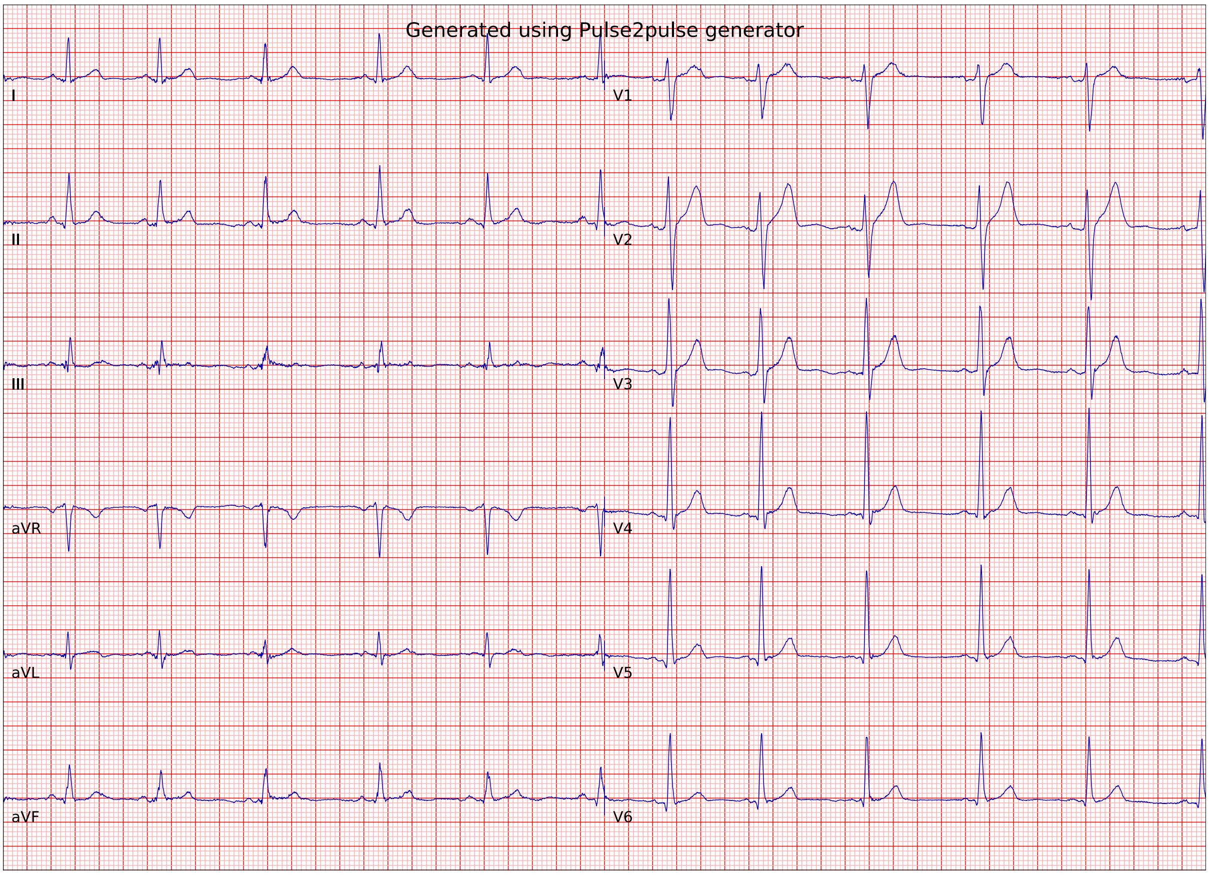
Generated using Pulse2pulse generator
I
II
III
aVR
aVL
aVF
V1
V2
V3
V4
V5
V6

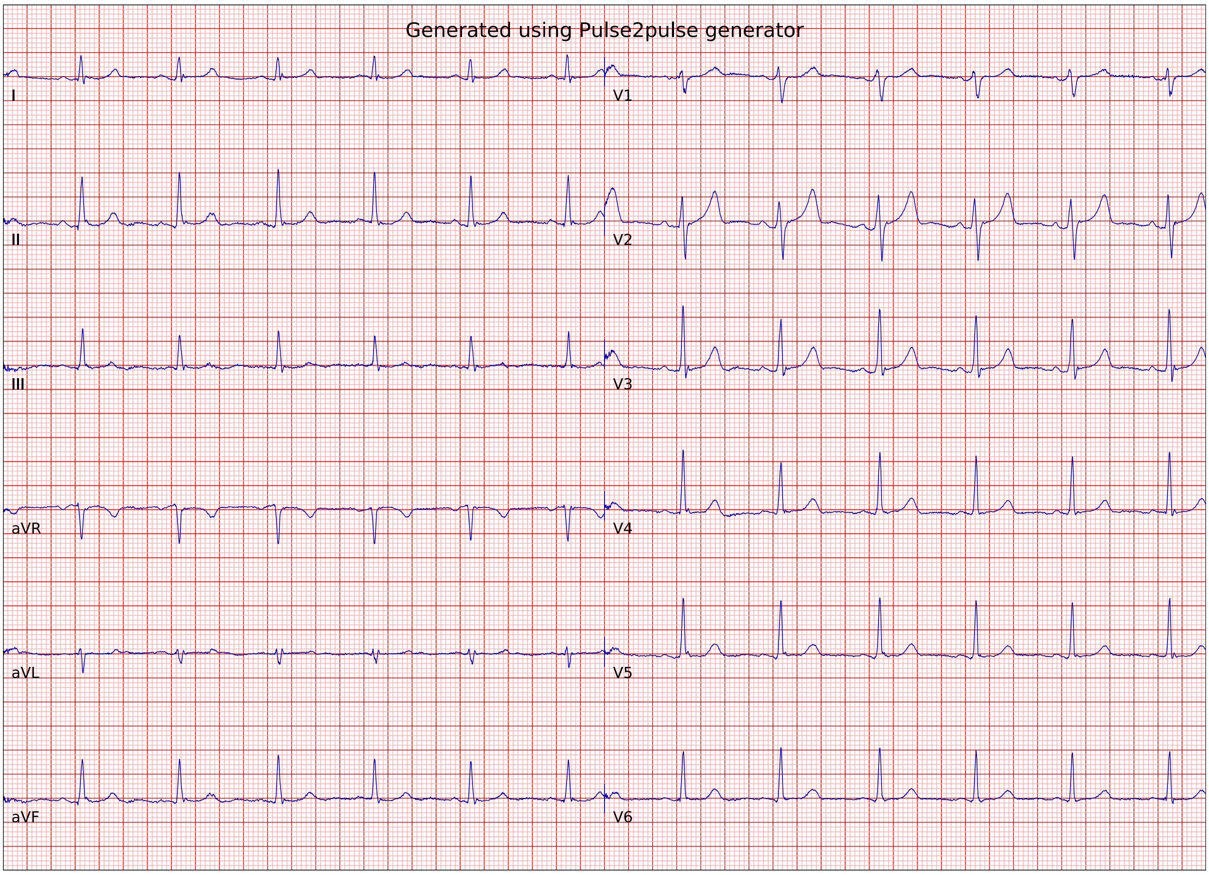
Generated using Pulse2pulse generator
I
II
III
aVR
aVL
aVF
V1
V2
V3
V4
V5
V6

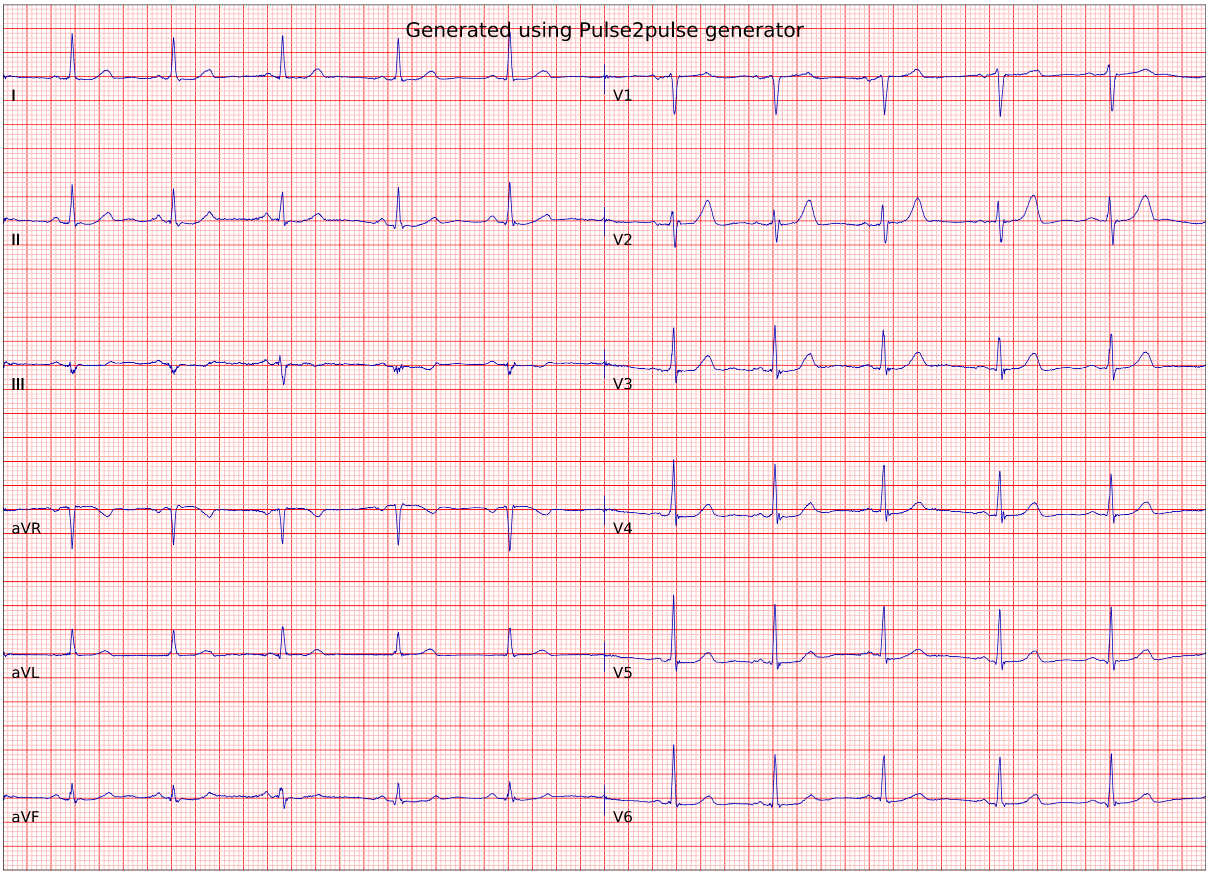
Generated using Pulse2pulse generator
I
II
III
aVR
aVL
aVF
V1
V2
V3
V4
V5
V6

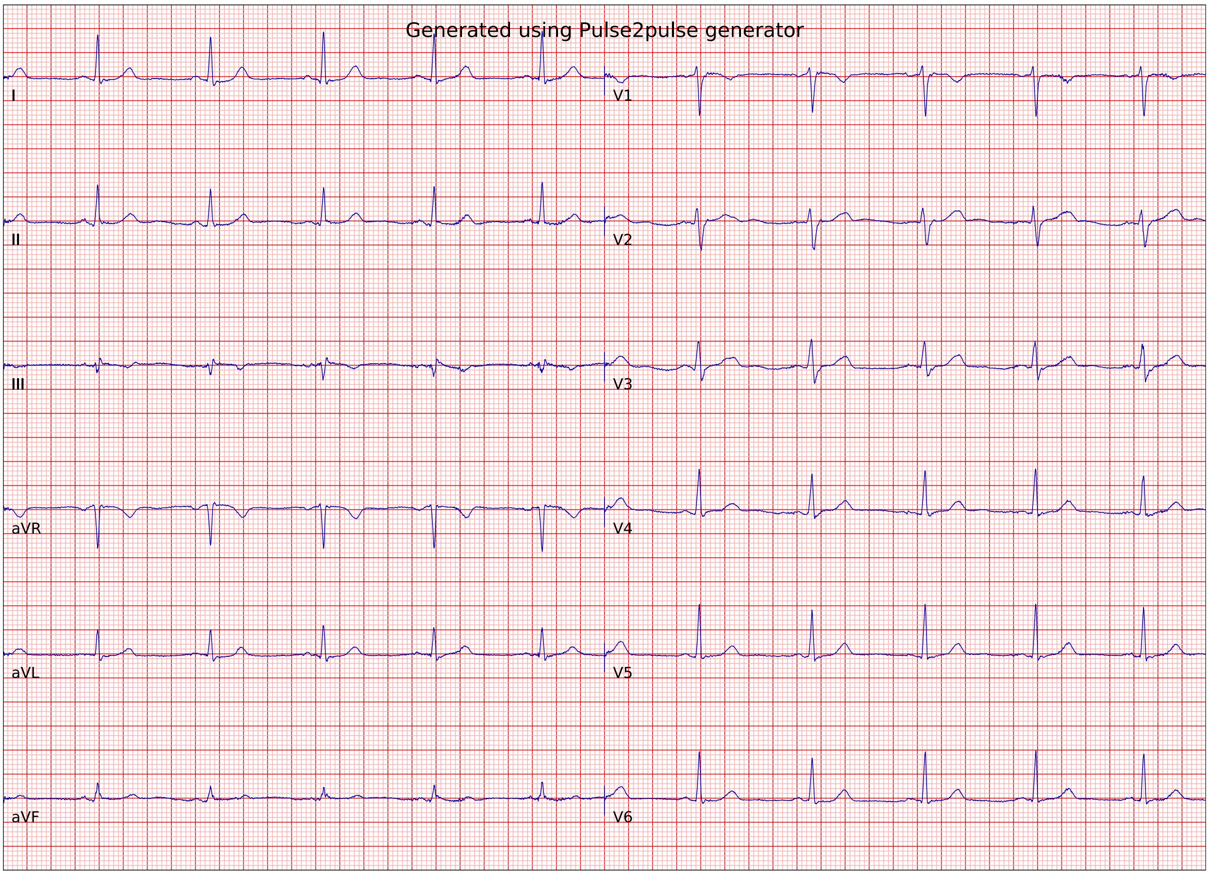
Generated using Pulse2pulse generator
I
II
III
aVR
aVL
aVF
V1
V2
V3
V4
V5
V6

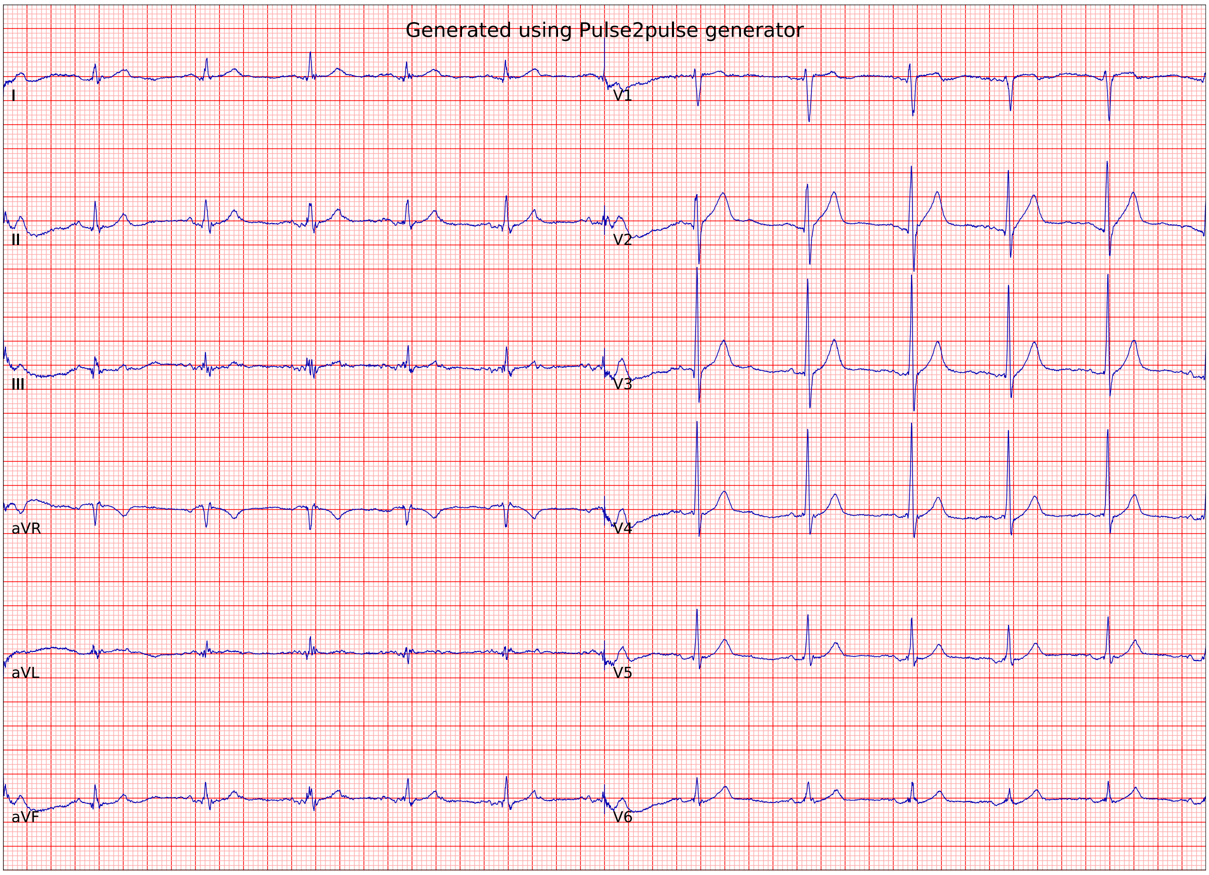
Generated using Pulse2pulse generator
I
II
III
aVR
aVL
aVF
V1
V2
V3
V4
V5
V6

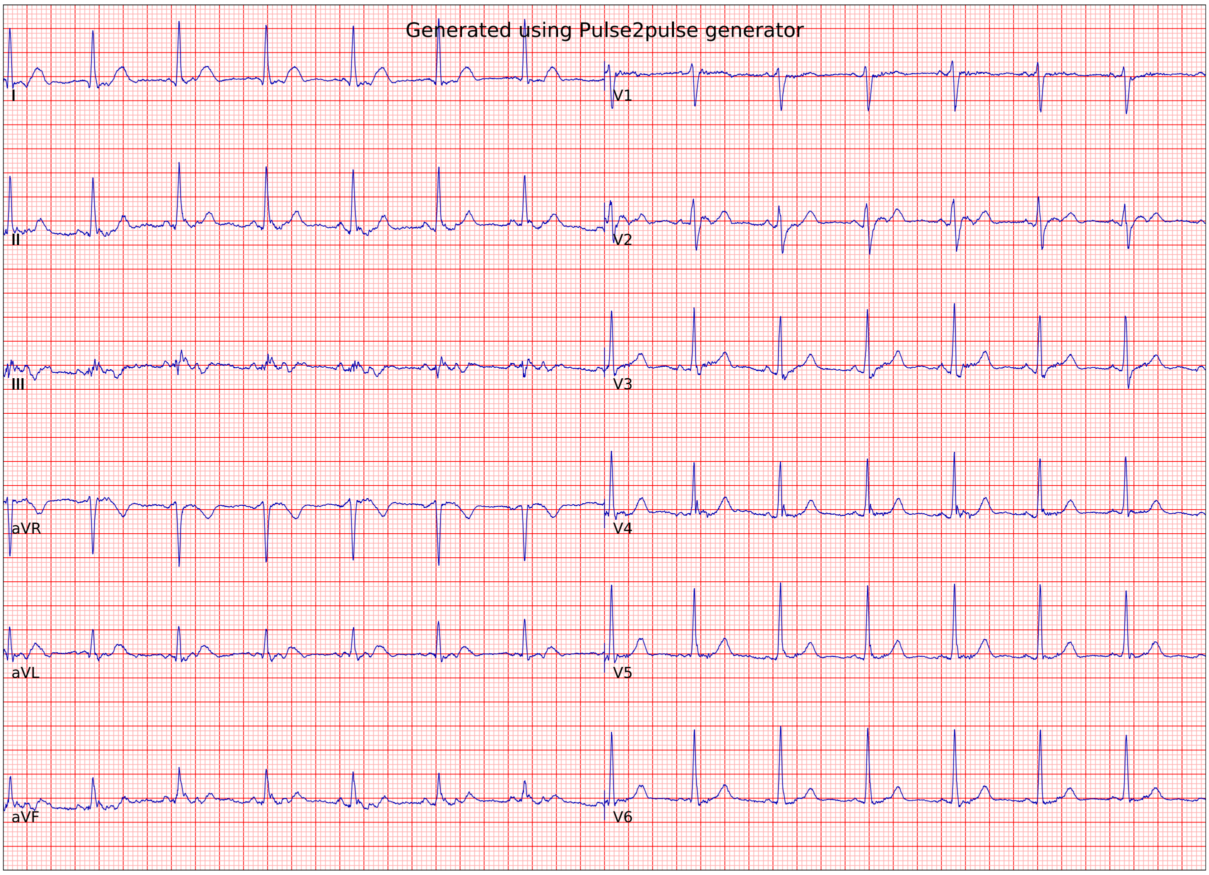
Generated using Pulse2pulse generator
I
II
III
aVR
aVL
aVF
V1
V2
V3
V4
V5
V6

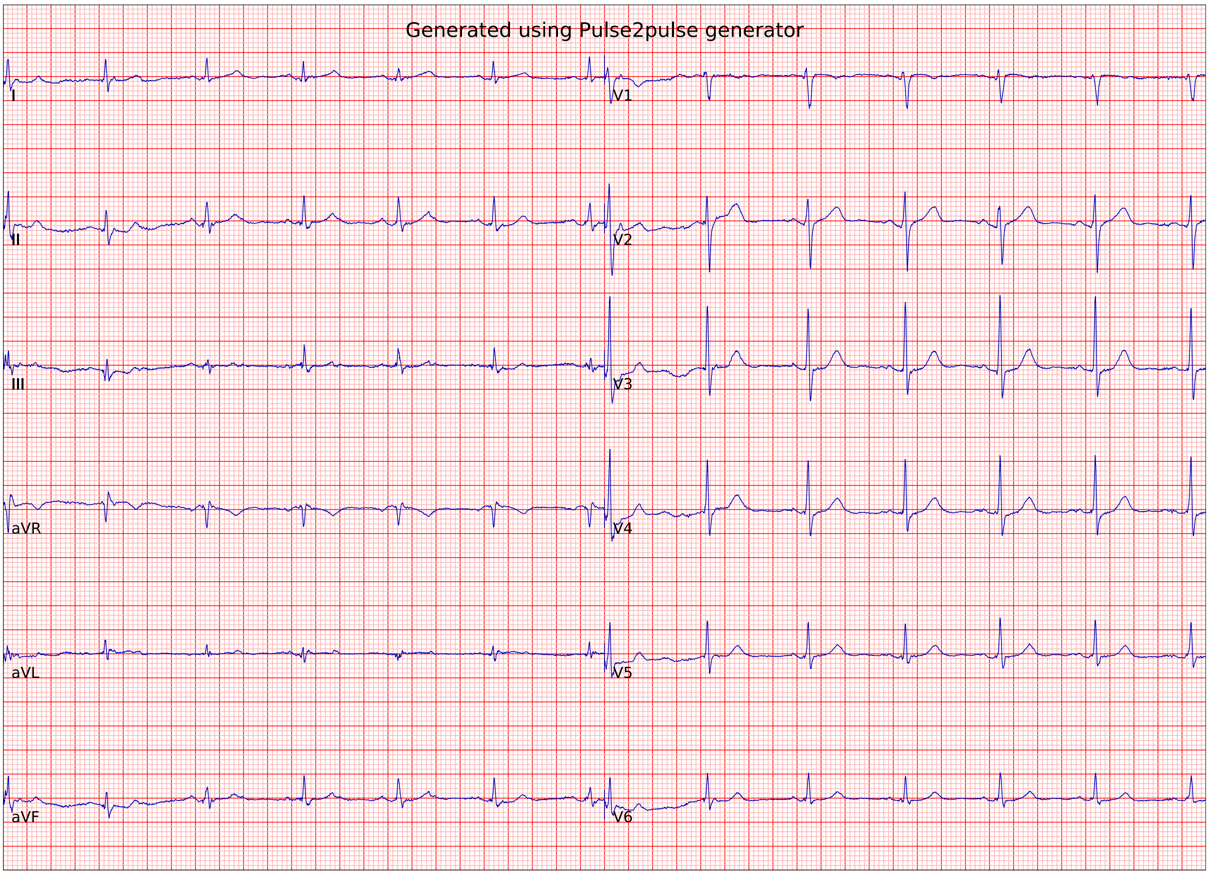
Generated using Pulse2pulse generator
I
II
III
aVR
aVL
aVF
V1
V2
V3
V4
V5
V6

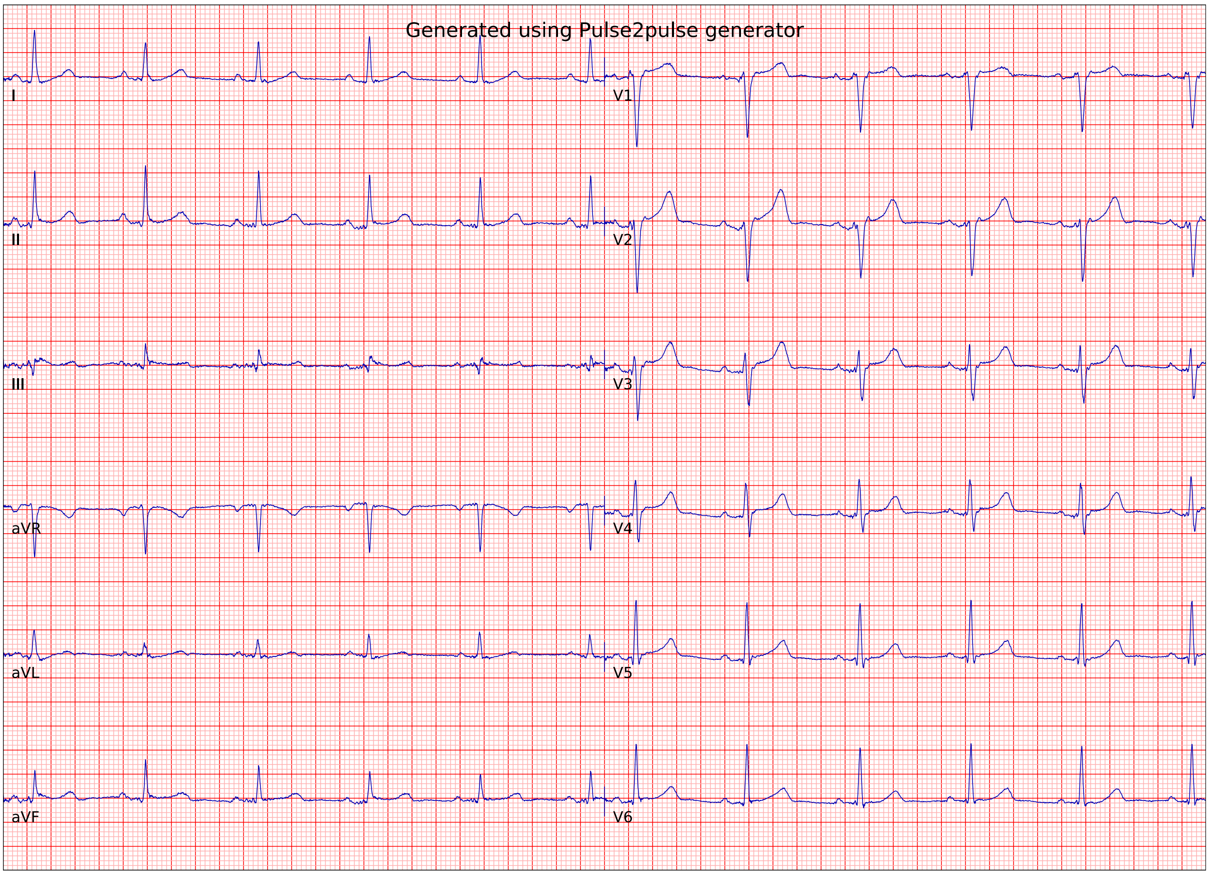
Generated using Pulse2pulse generator
I
II
III
aVR
aVL
aVF
V1
V2
V3
V4
V5
V6

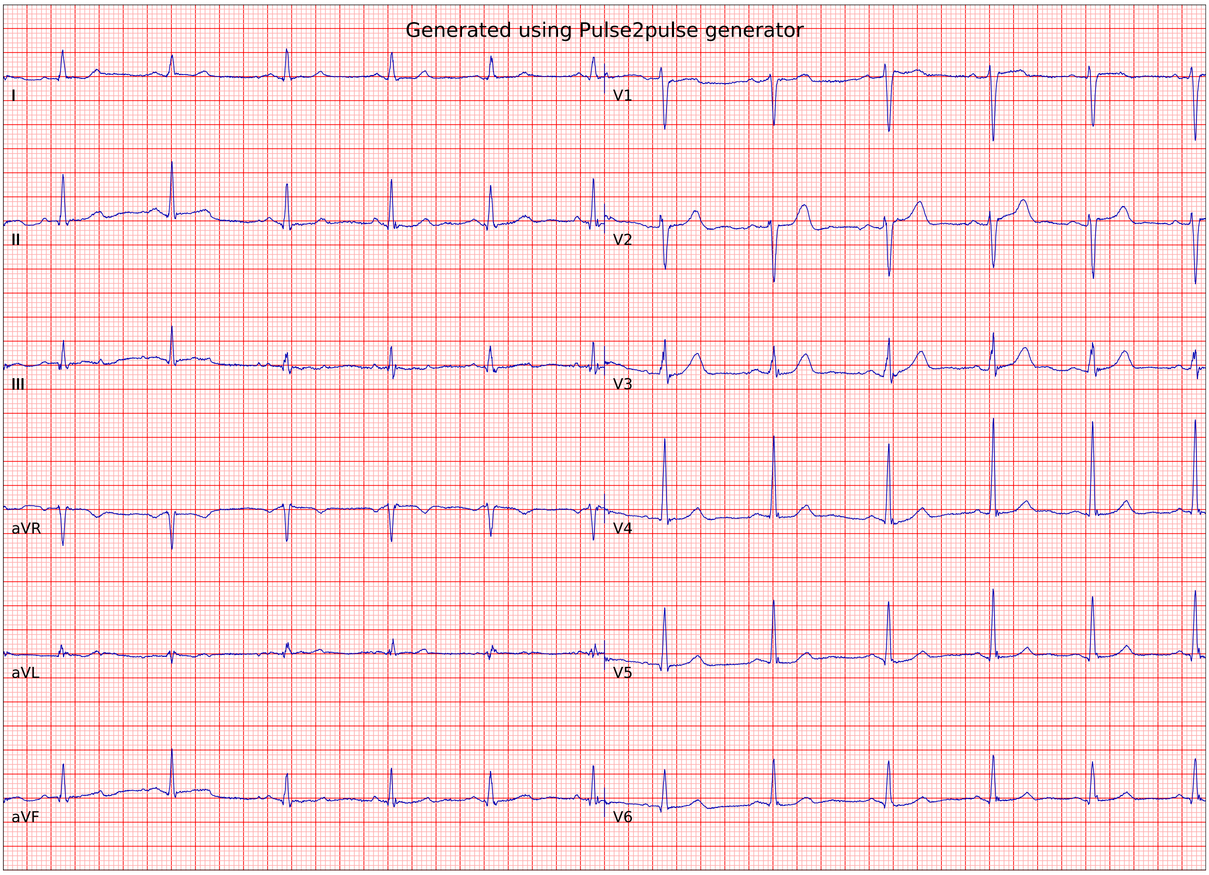
Generated using Pulse2pulse generator
I
II
III
aVR
aVL
aVF
V1
V2
V3
V4
V5
V6

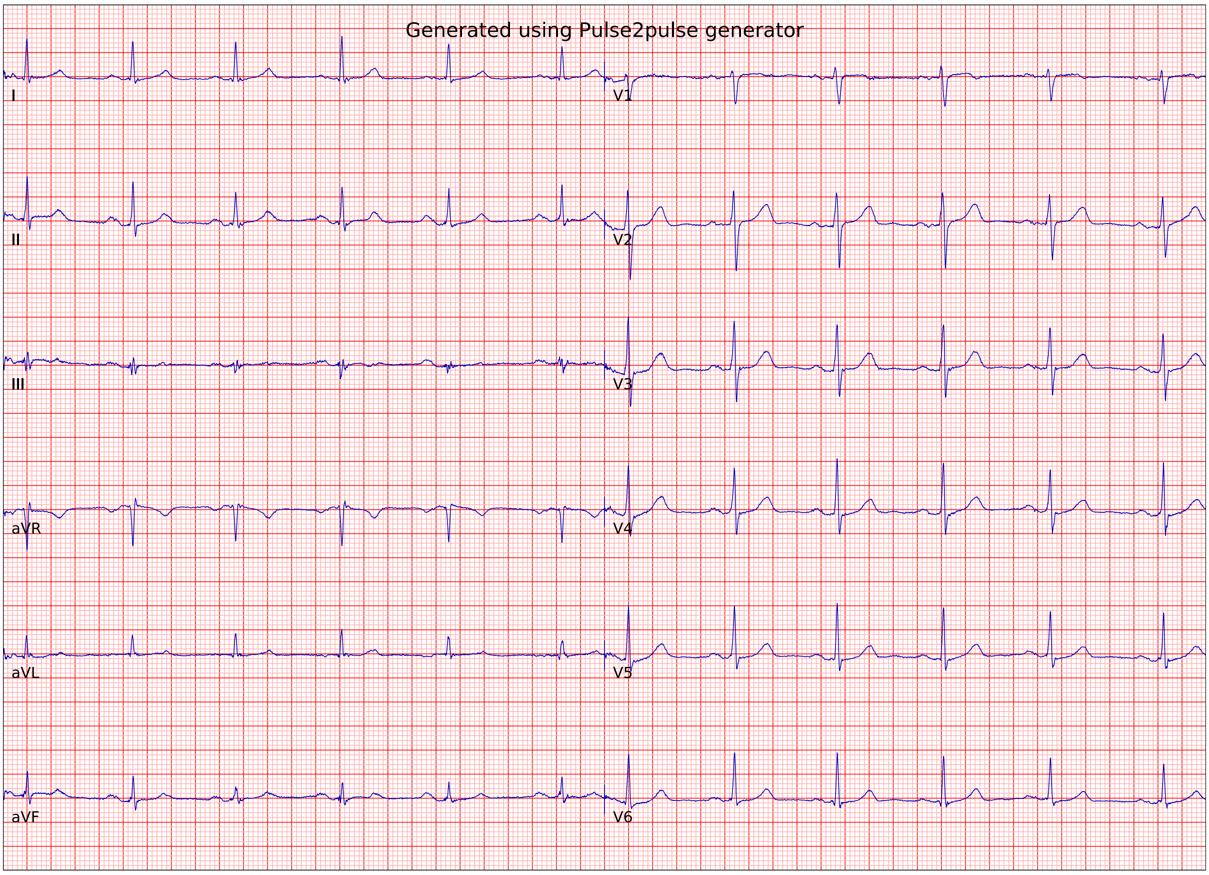
Generated using Pulse2pulse generator
I
II
III
aVR
aVL
aVF
V1
V2
V3
V4
V5
V6

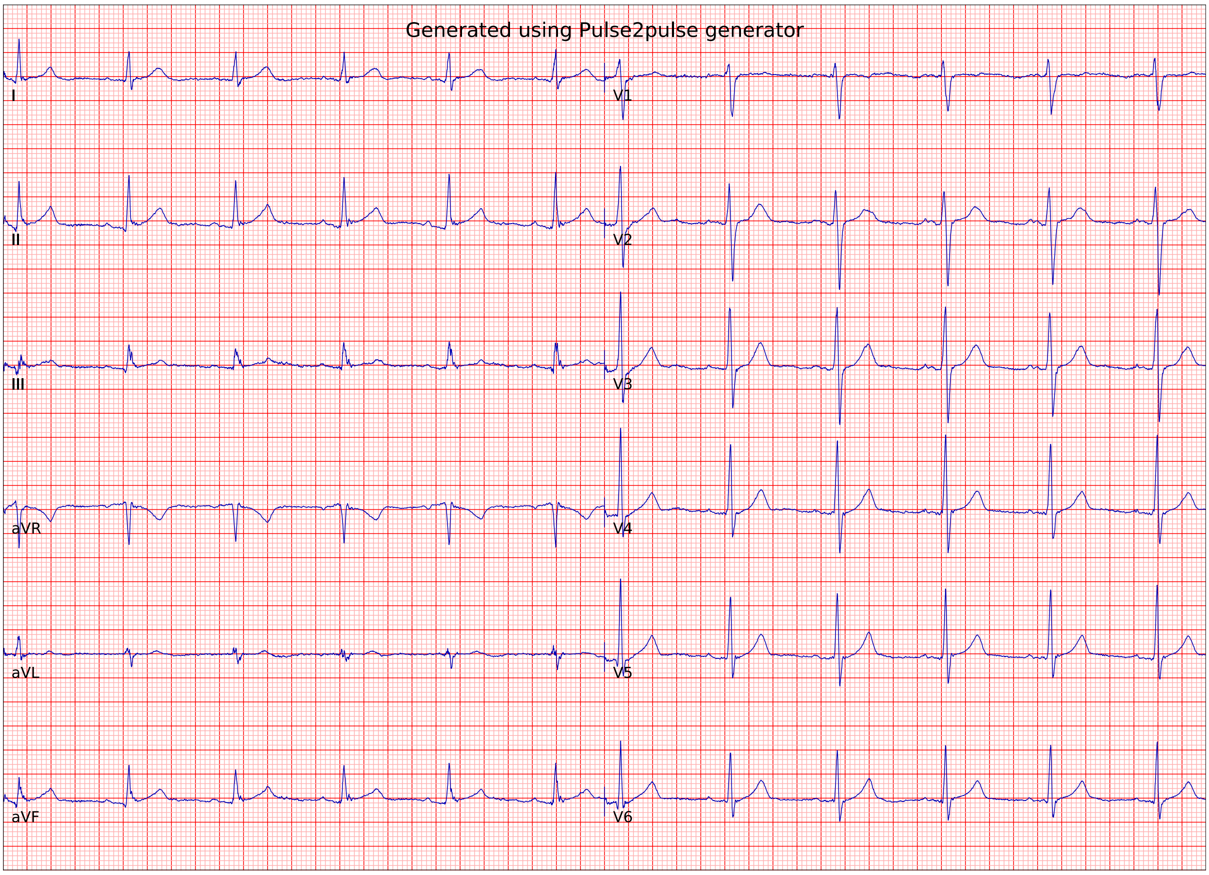
Generated using Pulse2pulse generator
I
II
III
aVR
aVL
aVF
V1
V2
V3
V4
V5
V6

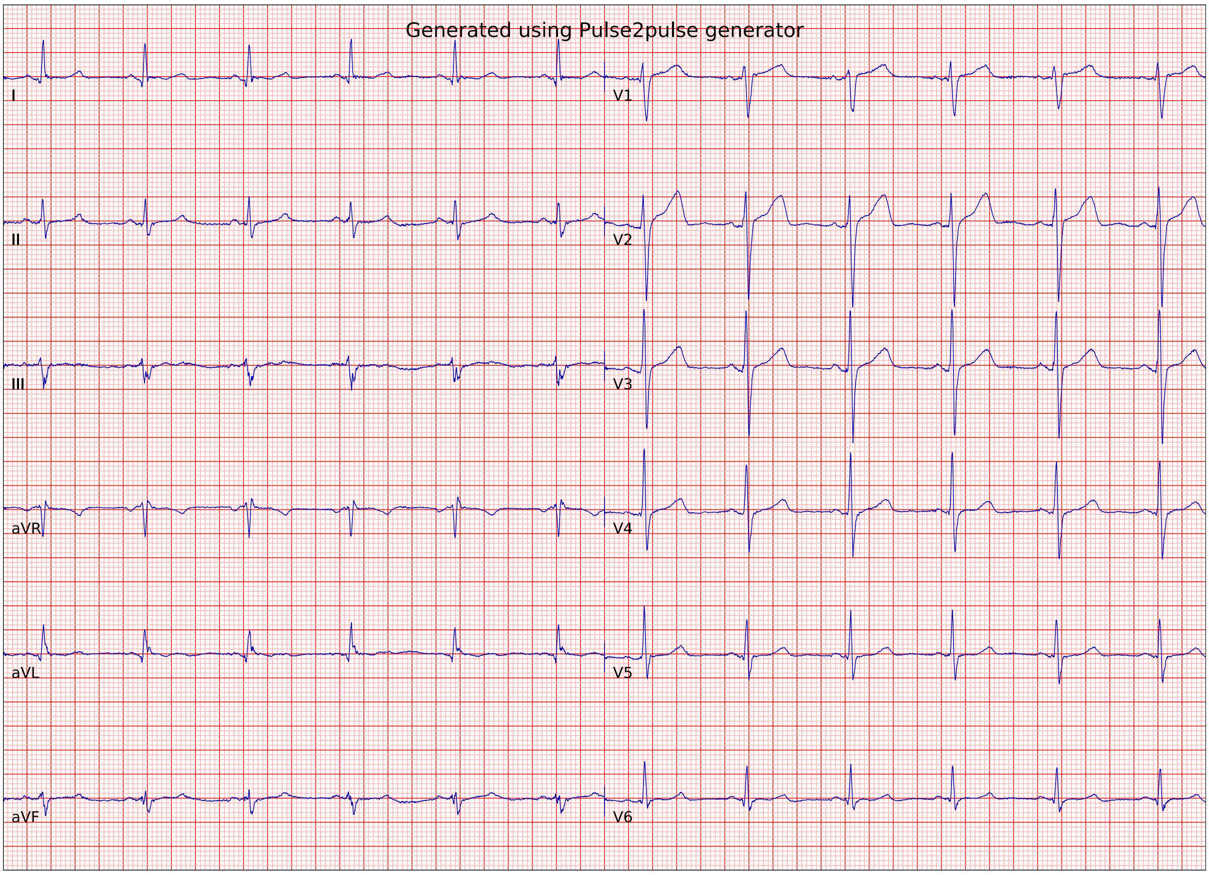
Generated using Pulse2pulse generator
I
II
III
aVR
aVL
aVF
V1
V2
V3
V4
V5
V6

**Figure S1**. Twenty randomly selected DeepFake ECGs. All the ECGs were selected from the Normal DeepFake ECGs.

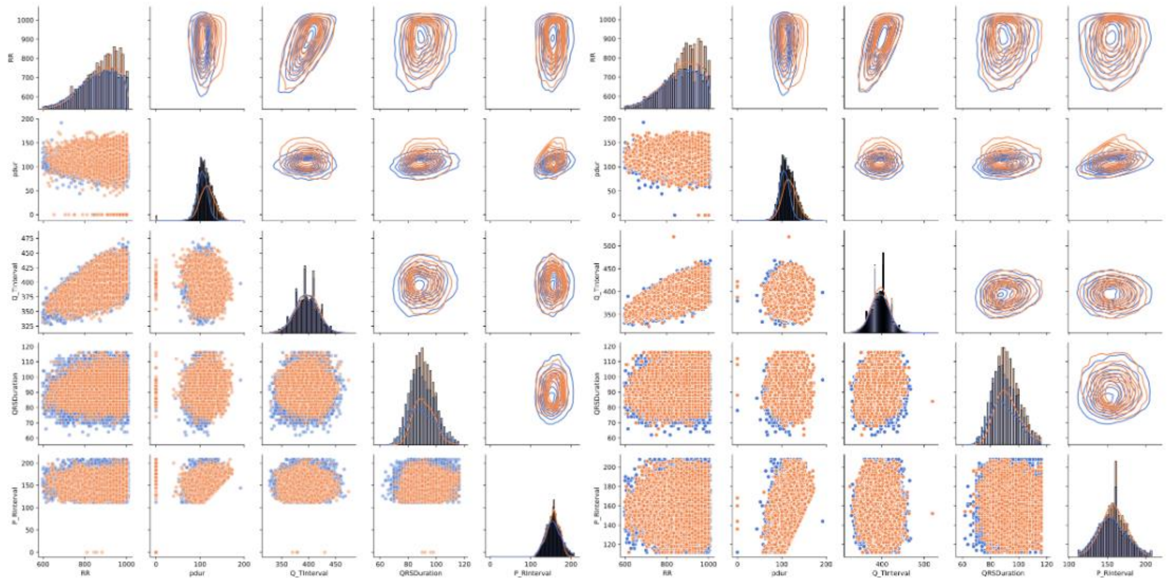


**(a)** From WaveGAN*.

**(b)** From Pulse2pulse.

**Figure S2**. The comparisons of the real and DeepFake distributions. Blue color plots represent real normal ECG distributions. Orange color plots represent the distribution of fake ECGs generated by Wavenet* (left lane (a)) and Pulse2pulse (Right Lane (b))
